# Surprisal-based large language models reveal immunologic insights in lobular breast cancer

**DOI:** 10.64898/2026.08.25.26361365

**Authors:** Bodhisattwa Prasad Majumder, Jessica Linak, Reece Adamson, Ruben Lozano Aguilera, Dhruv Agarwal, Zachary Reitz, Sophia Loiselle, Siddhartha Devarakonda, Peter Clark, Kelly Garneski Paulson, Sasha Stanton

## Abstract

In large data sets discovery is often limited to pre-conceived hypotheses and data fishing. Here we tested whether systematic exploration of AI generated hypotheses could uncover clinically meaningful signals in extensively studied data. We deployed AutoDiscovery, a newly launched large language model (LLM) framework designed to search for hypotheses based on surprisal and systematically interrogate complex datasets, on The Cancer Genome Atlas breast cancer cohort. The system did not identify clinically meaningful novel findings without human input. However, a seeded warm-start run with minimal text input from an oncologist revealed multiple interesting and surprising hypotheses. Among these was that a robust immune signature was present across all subtypes of invasive lobular carcinoma (ILC) that exceeded invasive ductal carcinoma (IDC). This observation was independently validated in independent cohorts and confirmed by high-sensitivity multi-immunofluorescence tumor tissue analyses. These results suggest immunotherapy approaches should be tested in ILC including early-stage ER+HER2-ILC; these patients are currently excluded from large neoadjuvant immunotherapy trials. They further demonstrate that surprisal-based hypothesis generation frameworks can extract previously unappreciated patterns from deeply interrogated cancer datasets and imply that disease domain experts working with LLMs can derive more meaningful insights from complex data than either could achieve alone.

## Introduction

Breast cancer is the most common malignancy in women and affects approximately one in eight women over their lifetime.^1^ Large-scale cancer profiling efforts have generated unprecedented datasets with the potential to transform our understanding of tumor biology and therapeutic response. Among these efforts, The Cancer Genome Atlas (TCGA) represented an investment of more than $1 billion across over 30 cancer types including invasive ductal carcinoma (IDC) and invasive lobular carcinoma (ILC) of the breast.^2 4^ These resources have yielded major biological insights, but their scale also contains inherent challenge; the volume and complexity of the data can obscure patterns. Due to this scale, analyses of large biomedical datasets often remain driven by the analysis of selected ‘fishing’ hypotheses. In this framework, data are interrogated with a predefined question which leaving substantial portions of the dataset unexplored. We hypothesized that systematic discovery-based approaches to heavily studied datasets could uncover additional clinically relevant insights in breast cancer. In particular, we were interested in insights for ILC. ILC is biologically distinct from IDC,^4^ with differences in detection, prognosis, and response to therapy,^5,6^ yet continues to be managed largely similarly in the clinic^7^ despite a paucity of lobular analysis in clinical trials. A systemic review of phase III/IV clinical trials for breast cancer has demonstrated that in 93 trials: ILC was excluded in 39 of the trials with only 13 trials documenting inclusion of ILC patients and 3 trials analysing ILC separately from IDC.^8^ For example despite early signals of improved response to pembrolizumab in metastatic PDL1+ ILC in Keynote 028, lobular pathology was an exclusion criteria in both of the neoadjuvant HR+HER2-trials Keynote 756 and Checkmate 7FL.^9,10^

Recent artificial intelligence (AI) approaches have demonstrated value in synthesizing and correlating biomedical knowledge. Large language model (LLM)–based tools can readily summarize evidence and assist with clinical documentation.^11,12^ However, these applications are descriptive: they test, organize, or restate prior knowledge rather than generate new hypotheses. Discovery in this setting requires a system designed not simply to retrieve but to identify unexpected, reproducible, and potentially meaningful patterns within complex datasets.

AutoDiscovery^13^ is an autonomous discovery-based framework that performs open-ended scientific exploration using Bayesian surprise. AutoDiscovery integrates LLMs with Monte Carlo tree search (MCTS) to perform open-ended hypothesis generation and evaluation without predefined research questions. The system is organized as a multi-agent architecture in which specialized LLM agents each perform a distinct role: a Hypothesis Generator that formulates testable scientific claims, an Experiment Generator that designs analysis plans, an Experiment Programmer that writes executable Python code, a Code Executor that runs verification procedures, an Experiment Analyst that interprets results, and an Experiment Reviewer and Reviser that validate and improve failed experiments. The tool makes both the analyses and the code immediately available for validation. Rather than evaluating all possible directions equally, AutoDiscovery prioritizes observations that are both surprising relative to prior expectations and reproducible across analyses, allowing iterative interrogation of the most informative signals. This allows for unbiased analyses of large data sets but focuses computational effort and hypothesis generation on the portions of the data most likely to yield biologically or clinically important findings. Here, we show that applying a surprisal-based large language model framework to TCGA breast cancer data, coupled with focused expert guidance, identifies new clinically actionable insights into ILC.

## Results

### Cold-start unguided surprisal-based analysis of breast cancer TCGA data

Autodiscovery^13^ analysis was performed previously acquired clinical and molecular data from patients with IDC and ILC ( 1,097) in the TCGA.^3,4^ Input features included clinical variables (for example, age, reported gender, race, and age at diagnosis), tumor characteristics (such as histological subtype, tissue source site, HER2 status, stage, and treatments), survival outcomes (including time to death and tumor event), somatic mutation data (available for 1,020 patients), and gene expression data (available for 1,068 patients). In an initial unguided ‘cold start’ analysis system was provided only basic information about the dataset structure (Methods), without hypotheses or input from a clinical expert. In this analysis, AutoDiscovery generated and analyzed 100 hypotheses, requiring a mean of 245 seconds per hypothesis (median, 231 seconds; range, 126–630 seconds) at an approximate cost of $0.19 USD per hypothesis. Of these, 89 were unique and 27 were classified as ‘positive’, indicating that the post-analysis result was more likely to be true than the pre-analysis expectation (**Fig. 1**). However, none was judged to be clinically useful or actionable.

**Figure 1.**
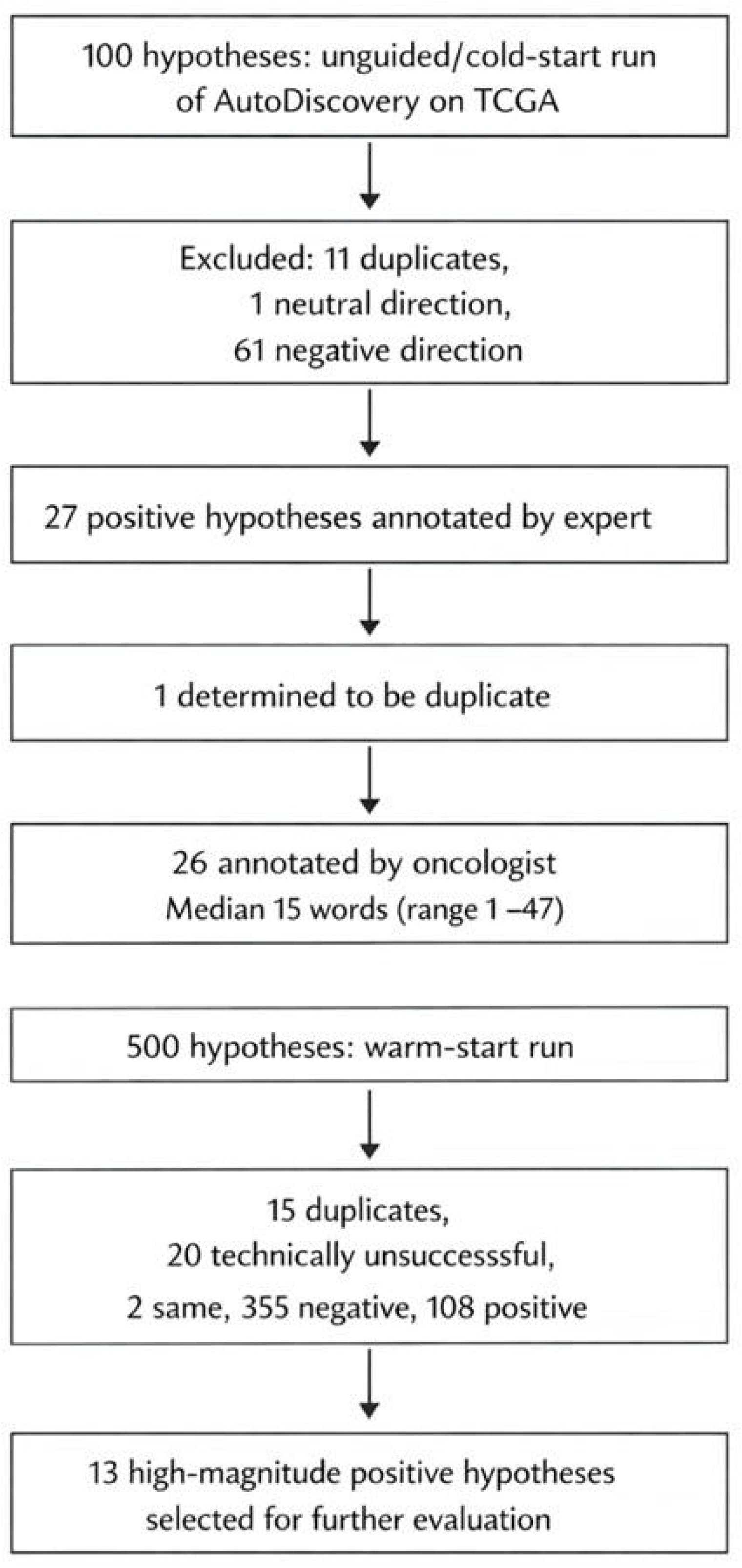
Experimental flow

### Expert-guided LLM discovery analysis reveals exhausted immune infiltrate in invasive lobular cancer

To improve on the limited clinical actionability of the initial unguided run, we hypothesized that iterative expert annotation would enhance the relevance of hypotheses generated by AutoDiscovery. A breast cancer physician-scientist with immunotherapy expertise annotated the 26 positive hypotheses from the first run, providing brief feedback on interpretive limitations or clinical usefulness. Annotation lengths ranged from 1 to 47 words per hypothesis (**Supplemental Table 1**); a 27th hypothesis was identified as a reworded duplicate that had not been removed during prior automated deduplication. Expert review required approximately 10 minutes per hypothesis, or ∼4 hours in total. We then performed a second, ‘warm-start’ AutoDiscovery run in which the system generated 500 hypotheses using both the original dataset structure information and the expert annotations (https://autodiscovery.allen.ai/runs/shared/swedish).

Among the 500 hypotheses generated and tested in the warm-start analysis, 15 were duplicates, 20 failed for technical reasons, and 108 were classified as positive. Many of these positive hypotheses reflected expected biology, including the highest post-analysis hypothesis implying enrichment of CDH1 alterations in invasive lobular carcinoma (ILC)^14^ Although such ‘true-positive’ findings supported the validity of the approach, they did not represent unexpected or clinically prioritizable observations. We therefore focused subsequent analyses on the 13 hypotheses with the largest positive shifts in belief, defined as an increase in posterior probability of more than 0.2 relative to the prior (**Supplemental Table 2**). The top two hypotheses in this group were notable but not immediately actionable. By contrast, the third-ranked hypothesis, “ILC tumors exhibit higher expression of the immune checkpoint genes PDCD1 and CD274 than IDC tumors after controlling for hormone receptor and HER2 status,” was both surprising and clinically consequential. This hypothesis shifted from a negative prior probability to a positive posterior probability, indicating that the analysis materially changed its estimated plausibility (**Fig. 2**). If validated, this observation would have important clinical implications, because ILC has historically been considered immunologically ‘cold’ and thus excluded from several immunotherapy trials, including KEYNOTE-756.^15^

**Figure 2.**
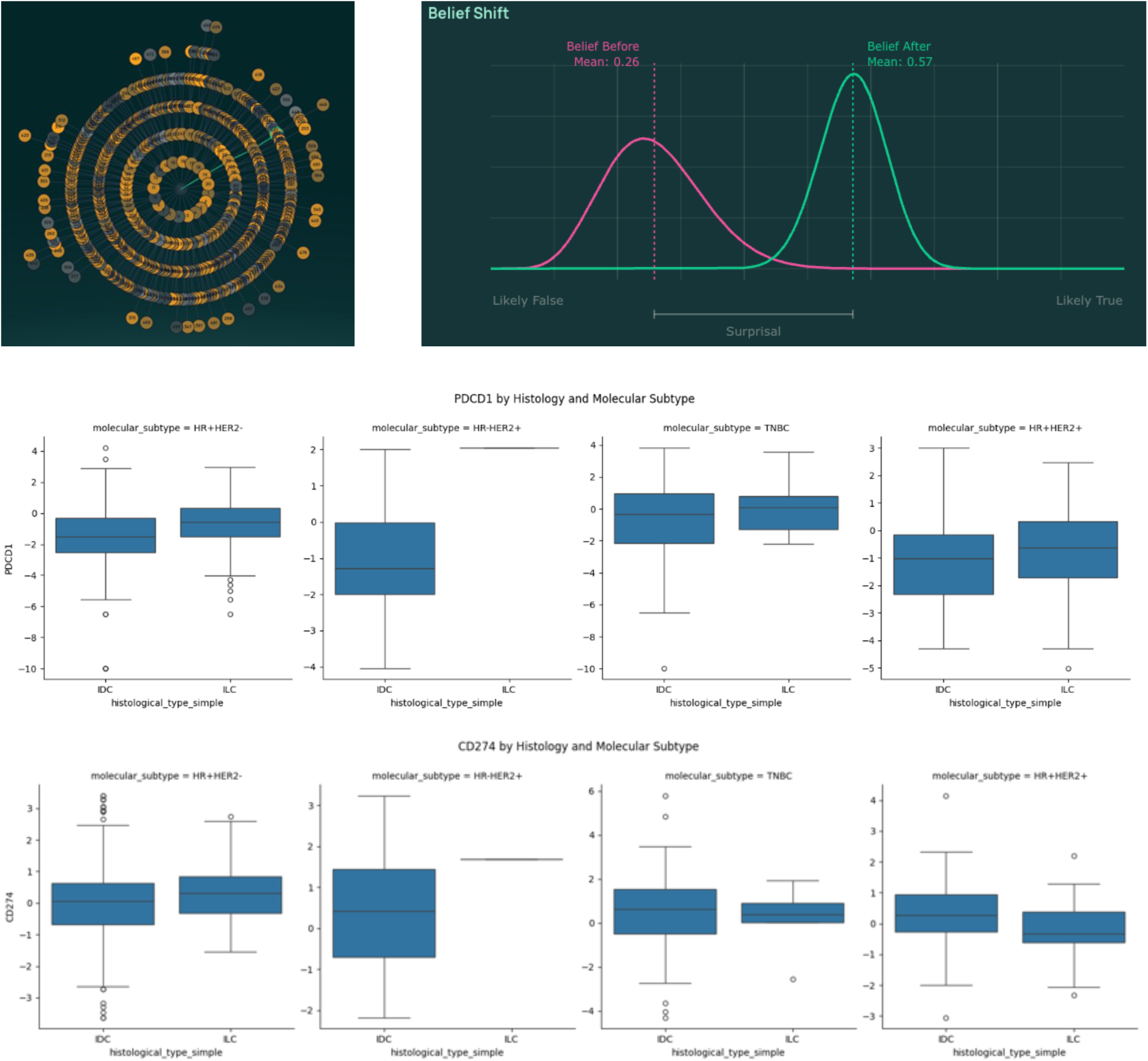
High expression of PDCD1 (PD-1) and CD274 (PD-L1) genes within ILC tumors. Top row: pre- and post-probability distributions for the AutoDiscovery generated hypotheses “Invasive lobular carcinoma (ILC) tumors exhibit higher expression of immune checkpoint genes PDCD1 and CD274 compared to invasive ductal carcinoma (IDC) tumors, after controlling for hormone receptor and HER2 status.” Middle row: analyses performed by AutoDiscovery to determine the post-probability for PDCD1, shown for key subgroups Bottom row: The same for CD274

### Validation of increased immune checkpoint expression in ILC in independent data set

To validate the increased expression of immune checkpoint genes in invasive lobular carcinoma (ILC), we next examined METABRIC, an independent large breast cancer cohort assembled in Europe.^16 18^ Consistent with the lower frequency of lobular relative to ductal carcinoma, fewer than 10% of the more than 2,000 breast cancers in this dataset were ILC (TCGA had specifically enriched for lobular). Owing to this limited sample size, we restricted the analysis to the largest subtype, estrogen receptor-positive, HER2-negative primary tumors, comprising 696 invasive ductal carcinomas (IDC) and 74 ILCs. Within this hormone receptor-positive, HER2-negative subgroup, ILC showed a modest but statistically significant increase in expression of both PDCD1 and CD274 relative to IDC. Median PDCD1 expression was 5.54 in ILC compared with 5.48 in IDC ( 0.034), whereas median CD274 expression was 5.51 in ILC compared with 5.45 in IDC ( 0.032) by Mann–Whitney U test (**Supplemental Methods**).^19^ This again supports hypothesis that a subset of ILCs may exist in a more immune-responsive state than previously appreciated.

### PD-1-positive T lymphocytes enriched in the ILC tumor microenvironment

We next turned to validate findings in ILC tumors. PD-L1 (CD274) protein expression in ILC by standard clinical immunohistochemistry (IHC) is reported to be rare;^20^ similarly, with canonical SP263 antibody only 3/12 (25%) ILC tumors we tested were PD-L1 positive with a combined positive (CPS) score >1% by and only 1/12 (8.3%) had a PD-L1 > 10%. However, the AutoDiscovery analyses suggested increased PDCD1 and CD274 RNA expression observed in ILC implying the presence of T lymphocytes and activation of the PD-1/PD-L1 checkpoint within the tumor microenvironment. We hypothesized that multiplex immunofluorescence, which could both detect PD-L1 expression at higher sensitivity and evaluate PD-1 and PD-L1 positive T lymphocytes, would better reveal checkpoint activation. Indeed, ILC tumors ( 12) harbored significantly greater numbers of PD-1-positive CD8-positive and CD4-positive T cells in the tumour microenvironment compared to normal breast tissue ( 6), a pattern that was recapitulated within the hormone receptor-positive, HER2-negative subgroup (**Fig. 3**). As expected, all 3 conventionally positive PD-L1 positive tumors had high PD-L1 positive T cell counts, but an additional 5 of 9 CPS negative tumors had brisk PD-L1 positive T cells in the ILC microenvironment thus supporting the findings from AutoDiscovery.

**Figure 3.**
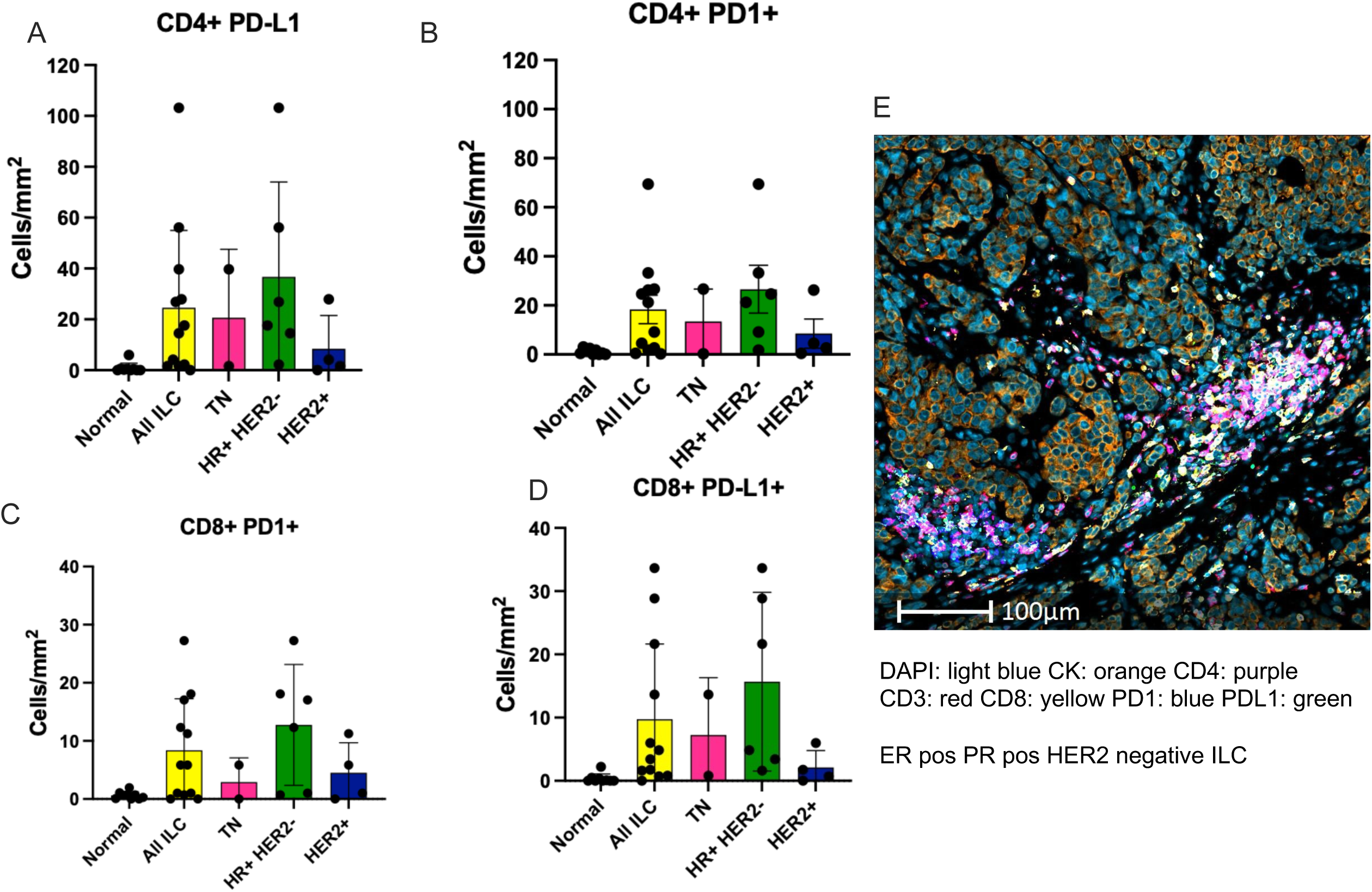
Enrichment of PD1+ and PDL1+ lymphocytes in the ILC microenvironment. ILC = invasive lobular carcinoma (n=12). Normal = healthy breast tissue (n=6). TN = triple negative. HR+ = hormone receptor positive. HER2 = her2. Panel E = example tumor.

## Discussion

Here we show that the autonomous hypothesis-generating LLM AutoDiscovery can extract clinically useful insights from extensively analyzed breast cancer datasets. Most notably, it identified the unexpected observation that invasive lobular carcinoma (ILC) harbors an immune-hot microenvironment enriched for PD-1-positive T cells across different breast cancer subtypes. This is important because anti-PD-1 immunotherapy is already approved for selected subsets of breast cancer^21,22^ and is under active investigation in others^15^, yet ILC has remained relatively understudied and has been excluded from several key early-stage trials, including KEYNOTE-756^15^. The expression of PDCD1 and CD274 implies exhausting infiltrating or neighboring lymphocytes. If a subset of ILC tumors is in fact immune-inflamed, this would provide a strong rationale for not excluding ILC from immunotherapy trials but instead specifically evaluating immunotherapy in this breast cancer context. Together, these data suggest that PD-1-directed and other immunotherapy-based clinical trials warrant broader evaluation in lobular breast cancers, including HR+ HER2-disease.

Given the importance of anti-PD1 immunotherapy across cancer types,^23^ there have been other explorations into role of immune response to ILC. Directed studies in TCGA have also suggested enhanced immune responses in ILC,^24^ our study builds on this by demonstrating this across ILC subtypes, using an unbiased approach, and validating in biological specimens. Other analyses have also supported surprisingly high ILC immune responses in ER+ HER2-disease. Post-hoc analyses of mammaprint testing on luminal ILC compared to luminal IDC showed significantly enhanced immune gene set enrichment in ILC compared to IDC, although different genes than in present study.^25^

There have also been early clinical explorations. Initial phase I studies of immune checkpoint inhibitors in advanced ER+ included ILC. For example in Keynote-028 trial of pembrolizumab in ER+ HER2-advanced breast cancer the ORR in ILC was 67% (2 of 3) as compared to 8% for IDC (1 of 13).^26^ The GELATO phase II study featured ILC alone and showed a clinical benefit rate of 26% of immune checkpoint inhibition combined with carboplatin low dose, there were responses across ILC subtypes supporting our findings.^27^ Importantly, in the present study we find brisk immune infiltrates across both early and late stage ILC including in the ER+ HER2-context.

Prior major published RCT of ICI in ER+ HER2-(Keynote 756) have demonstrated higher pathologic complete response rate with ICI addition but specifically excluded ILC in inclusion criteria.^15^ SWOG2206^28^ is similarly designed ongoing study, it includes but does not have stratification for IDC vs ILC. Our findings suggest that early-stage ILC is immune hot and that various immunotherapy strategies should be tested in these breast cancers.

This study has several limitations. Although the true-positive findings were broadly consistent with prior biological expectations, and the key result was independently supported by multiple approaches, we did not have the resources to validate every surprising hypothesis generated by the system. We anticipate some of the surprising hypotheses may be wrong, and all findings require downstream validation. In addition, AutoDiscovery did not operate in a fully unguided manner; performance was markedly improved by expert annotation rather than a true cold-start framework. This hybrid model is consistent with other emerging applications of LLMs in biomedicine, in which human expertise and machine inference together outperform either alone.^29,30^ The required level of expert input was feasible and achievable, with an estimated cost of approximately $0.20 per hypothesis evaluated and roughly 10 minutes of expert review per hypothesis.

Beyond direct immediate implications for ILC, this surprise-based autonomous discovery framework may provide a general strategy for time and cost-efficient extraction of new and valuable biological and clinical insights from existing datasets. The approach could be extended to other tumor types, to cohorts with richer therapeutic annotation, and to entirely different classes of biomedical data. Autonomous discovery systems, when combined with expert guided input and biologic secondary analyses, could become an important complement to biomedical investigation, helping to surface otherwise hidden, testable hypotheses

## Methods

### Ethics statement

All studies were IRB approved by Providence (IRB# Study2025000994-computational studies and Study2024000051-tissue analysis)

### Data sets

All cases of breast cancer available in the publicly available and previously anonymized cancer genome atlas (TCGA) were included,^3,4^ with all clinical, genomic, and gene expression data. For validation analyses, all breast cancer cases from METABRIC with ductal or lobular information were included.^16,17^

### Autodiscovery

AutoDiscovery^13^ was run using standard parameters. The initial run was a complete cold-start 100 hypothesis run where limited information was provided on data structure, the hypotheses were then individually annotated by oncologist and returned for a second warm-start run (**Supplemental methods**). For each sampled hypothesis, it was passed to the included multi-agent pipeline for experimental design, code generation, and execution. Generated Python code is run in a sandboxed environment with a 600-second timeout and with up to six debugging iterations permitted. Surprise is quantified as the difference between the mean of the prior and posterior belief distributions over the probability that a given hypothesis is supported by the data. Belief distributions are elicited before and after observing experimental results the LLM is sampled repeatedly for a Categorical judgment of hypothesis support from categories: Definitely True, Maybe True, Uncertain, Maybe False, Definitely False; the resulting response frequencies, where each category is represented by a scaler value: 1, 0.75, 05, 0.25, 0.0, define empirical distributions representing prior and posterior beliefs. Following search completion, hypotheses are deduplicated using LLM-based hierarchical agglomerative clustering (HAC). Hypotheses were then ranked by being positive hypotheses and strength of belief shift.

### DataVoyager

DataVoyager^31^ was applied to the TCGA and METABRIC breast cancer datasets to corroborate signals identified by AutoDiscovery. DataVoyager analyses were conducted in user-guided mode, with queries informed by the top-ranked AutoDiscovery hypotheses, enabling targeted re-examination of the immune checkpoint expression patterns in invasive lobular relative to invasive ductal carcinoma.

### PD-L1 immunohistochemistry

ILC tumors were stained with the SP263 antibody (Roche) under standard clinical conditions with combined positive score (CPS) performed by a clinical pathologist.

### Multicolor immunofluorescence

Please see Supplemental Methods. In brief, 21 formalin-fixed, paraffin-embedded (FFPE) breast tissue specimens (9 non-diseased breast and 12 ILC samples - 2 triple-negative, 4 HER2-positive, and 6 hormone receptor–positive/HER2-negative) were studied with Lunaphore COMET automated sequential immunofluorescence (Lunaphore Technologies, Tolochenaz, Switzerland). Slides were standardly prepared. For each sample, a 12.5 mm × 12.5 mm are was imaged at 20x magnification with autofluorescence followed by 15 iterative cycles of immunofluorescent staining, image acquisition, and signal elution, each with up to two primary antibodies. After completion of all cycles, a stacked multi-channel OME-TIFF file was imported into the HALO AI digital pathology software (version 4.2, Indica Labs, Albuquerque, NM). Using Mininet, two convolutional neural network classifiers were developed: one to mask imaging artifacts, and another to segment tissue into epithelial versus stromal regions. Cell nuclei were segmented in the DAPI channel using Nuclei Segmentation v2 algorithm (Halo AI, FL v1.0.0), with a 3 μm cytoplasmic expansion defining whole-cell boundaries. Cell phenotypes were assigned by applying intensity thresholding to nuclear and cytoplasmic signals for each biomarker. Cell counts were determined on a region-of-interest basis and normalized to tissue area. Data were analyzed using GraphPad Prism 10 (GraphPad Software, San Diego, CA). Differences among multiple groups were assessed using a Kruskal-Wallis test followed by Dunn’s multiple-comparisons test.^33^

## Data Availability

All data produced in the present work are publicly available, included in the manuscript, or available online at https://autodiscovery.allen.ai/runs/shared/swedish

## Code availability statement

Full code, analyses and results available at https://autodiscovery.allen.ai/runs/shared/swedish

## Acknowledgements

We thank Dr. Yaping Wu for reading the clinical PD-L1 IHC

## Author approval

All authors have seen and approved the manuscript

## References

1. Siegel, R. L., Kratzer, T. B., Wagle, N. S., Sung, H. & Jemal, A. Cancer statistics, 2026. CA Cancer J Clin 76, e70043 (2026).

2. Kandoth, C. et al. Mutational landscape and significance across 12 major cancer types. Nature 502, 333–339 (2013).

3. Cancer Genome Atlas Network. Comprehensive molecular portraits of human breast tumours. Nature 490, 61–70 (2012).

4. Ciriello, G. et al. Comprehensive Molecular Portraits of Invasive Lobular Breast Cancer. Cell 163, 506–519 (2015).

5. Pestalozzi, B. C. Clinical aspects of invasive lobular breast carcinoma (ILBC). Breast Dis 30, 1 (2008).

6. Chamalidou, C. et al. Survival patterns of invasive lobular and invasive ductal breast cancer in a large population-based cohort with two decades of follow up. Breast 59, 294–300 (2021).

7. Gradishar, W. J. et al. NCCN Guidelines® Insights: Breast Cancer, Version 5.2025. J Natl Compr Canc Netw 23, 426–436 (2025).

8. Baelen, K. V., Cauwenberge, J. V. & Maetens…, M. Reporting on invasive lobular breast cancer in clinical trials: a systematic review. … breast cancer (2024).

9. Loi, S., Salgado, R., Curigliano, G. & Diaz…, R. I. R. Neoadjuvant nivolumab and chemotherapy in early estrogen receptor-positive breast cancer: a randomized phase 3 trial. Nature medicine (2025).

10. Cardoso, F., O’Shaughnessy, J., Liu, Z. & McArthur…, H. Pembrolizumab and chemotherapy in high-risk, early-stage, ER^+^/HER2^−^ breast cancer: a randomized phase 3 trial. Nature Medicine (2025).

11. Parkinson, G. et al. Artificial Intelligence in Oncology: Clinical Applications, Challenges, and Opportunities. Am Soc Clin Oncol Educ Book 46, e520716 (2026).

12. Garrad, E. et al. Artificial Intelligence Among US Hematology Oncology Fellows: A Multicenter Survey of Education, Attitudes, and Clinical Use. JCO Oncol Pract OP2600433 (2026).

13. Agarwal, D. et al. Autodiscovery: Open-ended scientific discovery via bayesian surprise. Advances in Neural Information Processing Systems 38, 25181–25219 (2026).

14. Gamble, L. A. et al. Defining features of hereditary lobular breast cancer due to CDH1 with magnetic resonance imaging and tumor characteristics. NPJ Breast Cancer 9, 77 (2023).

15. Cardoso, F. et al. Pembrolizumab and chemotherapy in high-risk, early-stage, ER^+^/HER2^−^ breast cancer: a randomized phase 3 trial. Nat Med 31, 442–448 (2025).

16. Curtis, C. et al. The genomic and transcriptomic architecture of 2,000 breast tumours reveals novel subgroups. Nature 486, 346–352 (2012).

17. Pereira, B. et al. The somatic mutation profiles of 2,433 breast cancers refines their genomic and transcriptomic landscapes. Nat Commun 7, 11479 (2016).

18. Rueda, O. M. et al. Dynamics of breast-cancer relapse reveal late-recurring ER-positive genomic subgroups. Nature 567, 399–404 (2019).

19. Mann, H. B. & Whitney, D. R. On a test of whether one of two random variables is stochastically larger than the other. The annals of mathematical statistics (1947).

20. Shin, E., Kim, H. M. & Koo, J. S. Expression of PD-L1 in breast invasive lobular carcinoma. Plos one (2024).

21. Schmid, P. et al. Overall Survival with Pembrolizumab in Early-Stage Triple-Negative Breast Cancer. N Engl J Med 391, 1981–1991 (2024).

22. Schmid, P. et al. Pembrolizumab for Early Triple-Negative Breast Cancer. N Engl J Med 382, 810–821 (2020).

23. Ribas, A. & Wolchok, J. D. Cancer immunotherapy using checkpoint blockade. Science 359, 1350–1355 (2018).

24. Du, T. et al. Invasive lobular and ductal breast carcinoma differ in immune response, protein translation efficiency and metabolism. Sci Rep 8, 7205 (2018).

25. Lesnikoski, B. A., Crozier, J. A. & Srkalovic…, G. Abstract PS18-03: Differential gene expression in luminal-type invasive lobular carcinoma and invasive ductal carcinoma by MammaPrint risk stratification. Cancer … (2021).

26. Rugo, H. S. et al. Safety and Antitumor Activity of Pembrolizumab in Patients with Estrogen Receptor-Positive/Human Epidermal Growth Factor Receptor 2-Negative Advanced Breast Cancer. Clin Cancer Res 24, 2804–2811 (2018).

27. Voorwerk, L. et al. PD-L1 blockade in combination with carboplatin as immune induction in metastatic lobular breast cancer: the GELATO trial. Nat Cancer 4, 535–549 (2023).

28. Pusztai, L. Neoadjuvant chemotherapy and immunotherapy for estrogen receptor– positive human epidermal growth factor 2–negative breast cancer. Journal of Clinical Oncology (2024).

29. Zöller, N. et al. Human-AI collectives most accurately diagnose clinical vignettes. Proc Natl Acad Sci U S A 122, e2426153122 (2025).

30. Parikh, R. B. et al. Human-AI teaming to improve accuracy and efficiency of eligibility criteria prescreening for oncology trials: a randomized evaluation trial using retrospective electronic health records. Nat Commun 17, 2306 (2026).

31. Majumder, B. P., Surana, H., Agarwal, D. & Hazra…, S. Data-driven discovery with large generative models. arXiv preprint arXiv … (2024).

32. DuCote, T. J., Naughton, K. J., Skaggs, E. M. & Bocklage…, T. J. Using artificial intelligence to identify tumor microenvironment heterogeneity in non–small cell lung cancers. Laboratory … (2023).

33. Kruskal, W. H. & Wallis, W. A. Use of ranks in one-criterion variance analysis. Journal of the American statistical Association 47, 583–621 (1952).

