## Supplemental Methods for "Surprisal-based large language models reveal immunologic insights in lobular breast cancer"

Majumder et al

**First Run – Autodiscovery**

TCGA Breast Cancer Metadata -

**Overview**

| **Domain** | cancer genomics |
| --- | --- |
| **Workflow Tags** | clinical data analysis |

**Datasets**

**cleaned_clinical_data.csv**

Clinical and demographic metadata for TCGA breast cancer cohort, including diagnosis, treatment, outcomes, and patient characteristics.

**Columns**

| **Column Name** | **Description** |
| --- | --- |
| bcr_patient_barcode | TCGA patient barcode (BCR identifier) |
| gender | Reported gender |
| race | Race (list/label) |
| ethnicity | Ethnicity |
| days_to_birth | Negative days offset from birth |
| age_at_diagnosis | Age at initial pathologic diagnosis (years) |
| menopause_status | Menopause status at diagnosis |
| histological_type | Histological tumor type |
| diagnosis_year | Year of initial pathologic diagnosis |
| icd_10_code | ICD-10 disease classification code |
| tissue_source_site | Tissue source site code |
| prior_malignancy | History of prior malignancy diagnosis |
| er_status | Estrogen receptor status (Positive/Negative/Indeterminate) |
| pr_status | Progesterone receptor status (Positive/Negative/Indeterminate) |
| her2_ihc_status | HER2 immunohistochemistry receptor status |
| her2_fish_status | HER2 FISH (fluorescence in situ hybridization) result |
| her2_ihc_score | HER2 IHC score (0, 1+, 2+, 3+) |
| pathologic_stage | AJCC pathologic stage (post-surgery) |
| pathologic_t | AJCC pathologic T (tumor) stage |
| pathologic_n | AJCC pathologic N (lymph node) stage |
| pathologic_m | AJCC pathologic M (metastasis) stage |
| lymph_nodes_examined | Number of lymph nodes examined |
| lymph_nodes_positive | Number of lymph nodes positive by H&E |
| anatomic_subdivision | Anatomic neoplasm subdivision (breast quadrant/laterality) |
| tumor_status | Person neoplasm cancer status (tumor free/with tumor) |
| surgery_type | Breast carcinoma surgical procedure name |
| surgical_margin | Surgical margin status (Positive/Negative/Close) |
| neoadjuvant_treatment | History of neoadjuvant treatment |
| radiation_therapy | Radiation therapy administered (YES/NO) |
| pharmaceutical_therapy | Postoperative pharmaceutical therapy administered |
| drug_name | Primary drug name (first occurrence) |
| therapy_type | Primary therapy type (first occurrence) |
| vital_status | Patient vital status (Alive/Dead) |
| days_to_last_followup | Days from diagnosis to last follow-up |
| days_to_death | Days from diagnosis to death |
| new_tumor_event | New tumor event after initial treatment (YES/NO) |
| days_to_new_tumor | Days to new tumor event after initial treatment |

**cleaned_gene_expression.csv**

**dataset_info:**

name: TCGA-BRCA Gene Expression (RNA-Seq)

source: UCSC Xena - TOIL RSEM FPKM

normalization: RSEM expected_count, then log2(fpkm+0.001) transformed

platform: Illumina HiSeq RNA-Seq

data_type: Gene expression quantification

url: https://toil.xenahubs.net/download/tcga_RSEM_gene_fpkm.gz

**dimensions:**

genes: 60498

samples: 1068

description: Rows are genes (Ensembl gene IDs), columns are samples (TCGA patient

barcodes)

**sample_alignment:**

clinical_patients: 1068

matched_samples: 1068

note: Samples filtered to match patients in cleaned_clinical_data.csv

**data_description:**

values: Log2(FPKM + 0.001) - normalized gene expression levels

interpretation: Higher values indicate higher gene expression. Values are continuous

and log-transformed.

missing_data: Genes with all zeros or NaN across samples were removed

**usage_notes:**

- Gene identifiers are in the index (first column)

- Sample identifiers are in the column headers (TCGA patient barcodes)

- Data is pre-normalized and log-transformed, ready for analysis

- Match samples to clinical data using patient barcode (column headers)

- For patient-level analysis, data is already aggregated to one sample per patient

**cleaned_mutations.csv**

**dataset_info:**

name: TCGA-BRCA Somatic Mutations

source: GDC Data Portal - MC3 Public MAF

format: Mutation Annotation Format (MAF)

data_type: Somatic mutations (SNVs and small indels)

workflow: MC3 - Multi-Center Mutation Calling in Multiple Cancers

description: Curated somatic mutation calls combining multiple variant calling pipelines

**dimensions:**

total_mutations: 135030

unique_patients: 1020

unique_genes: 18907

columns: 21

**sample_alignment:**

clinical_patients: 1097

patients_with_mutations: 1020

note: Mutations filtered to match patients in cleaned_clinical_data.csv

**variant_summary:**

top_variant_classifications:

Missense_Mutation: 69328

Silent: 25786

3'UTR: 9585

Frame_Shift_Del: 9324

Nonsense_Mutation: 5890

Intron: 4617

5'UTR: 3484

Splice_Site: 2084

RNA: 1728

Frame_Shift_Ins: 961

3'Flank: 721

In_Frame_Del: 683

5'Flank: 606

Translation_Start_Site: 93

Nonstop_Mutation: 90

In_Frame_Ins: 50

variant_types:

SNP: 122135

DEL: 11564

INS: 1326

ONP: 5

description: Most common mutation types in the dataset

**top_mutated_genes:**

genes:

TTN: 446

PIK3CA: 425

TP53: 376

MUC16: 213

CDH1: 144

MAP3K1: 142

GATA3: 141

KMT2C: 132

RYR2: 121

SYNE1: 118

FLG: 107

DST: 106

HMCN1: 103

DMD: 92

NEB: 91

OBSCN: 91

USH2A: 88

RYR3: 85

SPTA1: 84

SYNE2: 80

description: Top 20 most frequently mutated genes

**column_descriptions:**

Hugo_Symbol: HUGO gene symbol

Chromosome: Chromosome where mutation occurs

Start_Position: Genomic start position (1-based)

Variant_Classification: Mutation type (e.g., Missense_Mutation, Nonsense_Mutation)

Variant_Type: SNP, DNP, INS, or DEL

HGVSp_Short: Protein change annotation

Tumor_Sample_Barcode: TCGA sample barcode

bcr_patient_barcode: Patient identifier (TCGA-XX-XXXX format)

**usage_notes:**

- Each row represents one somatic mutation

- Match to clinical data using the bcr_patient_barcode column

- For gene-level analysis, aggregate mutations by Hugo_Symbol

- For patient-level mutation burden, count mutations per bcr_patient_barcode

- Variant_Classification indicates functional impact

- FILTER column shows variant quality filtering status

**Columns**

| **Column Name** | **Description** |
| --- | --- |
| Hugo_Symbol | HUGO gene symbol |
| Chromosome | Chromosome where mutation occurs |
| Start_Position | Genomic start position (1-based coordinate) |
| End_Position | Genomic end position (1-based coordinate) |
| Strand | DNA strand on which the mutation is annotated (+ or -) |
| Variant_Classification | Functional class of the mutation (e.g., Missense_Mutation, Nonsense_Mutation, RNA, UTR, Frame_Shift) |
| Variant_Type | Type of variant call (e.g., SNP, DEL, INS, ONP) |
| Reference_Allele | Reference allele observed at the mutation site |
| Tumor_Seq_Allele1 | First observed tumor allele at the locus |
| Tumor_Seq_Allele2 | Second observed tumor allele at the locus |
| Tumor_Sample_Barcode | TCGA tumor sample barcode |
| bcr_patient_barcode | Patient identifier (TCGA-XX-XXXX format) |
| HGVSc | HGVS coding DNA change annotation (c.) using transcript coordinates |
| HGVSp | HGVS protein change annotation using full protein notation (p.) |
| HGVSp_Short | Shortened HGVS protein change notation |
| Transcript_ID | Ensembl transcript identifier (ENST...) used for annotation |
| Exon_Number | Exon where the variant occurs (e.g., 6/6) |
| IMPACT | Predicted functional impact class of the mutation (HIGH, MODERATE, LOW, MODIFIER) |
| SIFT | SIFT protein impact prediction in the form label(score), where label is one of tolerated/deleterious/deleterious_low_confidence and score is a probability (0–1) |
| PolyPhen | PolyPhen-2 protein impact prediction in the form label(score), where label is one of benign/possibly_damaging/probably_damaging and score is a probability (0–1) |
| FILTER | Variant filter status indicating whether the call passed QC filtering |

### Run Configuration (args.json)

| **Parameter** | **Value** |
| --- | --- |
| dataset_metadata | s3://ai2-asta-workspaces/autods/datasets/tcga-breast-cancer/tcga_breast_cancer_metadata.json |
| out_dir | /results |
| model | o4-mini |
| belief_model | o4-mini |
| user_query | null |
| temperature | null |
| belief_temperature | null |
| reasoning_effort | medium |
| continue_from_dir | null |
| continue_from_json | null |
| n_experiments | 100 |
| k_experiments | 8 |
| allow_generate_experiments | True |
| n_belief_samples | 30 |
| timestamp_dir | True |
| exploration_weight | 2.0 |
| dataset_metadata_type | dbench |
| work_dir | work |
| delete_work_dir | True |
| beam_width | 8 |
| use_beam_search | False |
| mcts_selection | ucb1_recursive |
| pw_k | 1.0 |
| pw_alpha | 0.5 |
| k_parents | 3 |
| implicit_bayes_posterior | False |
| surprisal_width | 0.2 |
| belief_mode | boolean_cat |
| use_binary_reward | False |
| dedupe | True |
| only_save_results | False |
| experiment_first | False |
| code_timeout | 1800 |
| run_eda | False |
| n_warmstart | 8 |
| use_online_beliefs | False |
| evidence_weight | 2.0 |
| kl_scale | 5.0 |
| reward_mode | kl |
| warmstart_experiments | null |

**Best Run - Autodiscovery**

TCGA Breast Cancer Metadata (Notes)

### Overview

| **ID** | 0 |
| --- | --- |
| **Domain** | cancer genomics |
| **Workflow Tags** | clinical data analysis |

### Datasets

#### cleaned_clinical_data.csv

**Clinical and demographic metadata for TCGA breast cancer cohort, including diagnosis, treatment, outcomes, and patient characteristics. Insight from prior experiments:**

- Hypothesis: Among patients with PIK3CA mutations, there is a characteristic co-mutation pattern wherein mutations in TP53 occur less frequently than expected by chance. Oncologist Comment: unexpected, not looked at per literature. Would evaluate Lobular and ductal diffferently as TP53 has different expression and prognosis between them.

-Hypothesis: Patients treated with lumpectomy have different overall survival distributions compared to those treated with mastectomy, after controlling for pathologic stage. Oncologist Comment: Would only evaluate lumpectomy vs mastectomy for stage I-III patients, not done in stage IV. Would include I-III as might differ in more advanced patients

- Hypothesis: Higher tumor mutational burden is associated with decreased gene expression heterogeneity, as measured by per-sample expression standard deviation. Oncologist Comment: expected

- Hypothesis: African American breast cancer patients exhibit a higher overall somatic mutation burden than White patients. Oncologist Comment: would separate HR+, HER2+, and TNBC. Suspect it is because more TNBC in AA patients

- Hypothesis: African American breast cancer patients have a higher tumor mutational burden (TMB) than White patients. Oncologist Comment: would separate HR+, HER2+, and TNBC. Suspect it is because more TNBC in AA patients

- Hypothesis: Patients with HIGH‐impact TP53 mutations have shorter overall survival than those with low/moderate‐impact or TP53 wild-type, controlling for stage and age. Oncologist Comment: expected, would separate out stage I-III (unknown) and IV (known) also would look at lobular vs ductal.

- Hypothesis: Hispanic or Latino ethnicity is associated with a different tumor mutational burden compared to non-Hispanic patients. Oncologist Comment: would not expect hispanic patients have higher TMB so expected.

- Hypothesis: Triple-negative breast cancer patients (ER–, PR–, HER2–) exhibit higher tumor mutational burden than non–triple-negative patients. Oncologist Comment: expected, TMB can be higher in TNBC but depends on BRCA mutation carrier and would not expect across all TNBC.

- Hypothesis: Patients with basal-like subtype (ER−, PR−, HER2−) exhibit higher immune checkpoint gene expression than luminal subtypes. Oncologist Comment: expected, ICI expression can be seen in both subtypes

- Hypothesis: Patients with frameshift mutations in core DNA damage repair (DDR) genes (BRCA1, BRCA2, ATM, TP53) have higher overall tumor mutational burden compared to patients without frameshift mutations in these genes. Oncologist Comment: unexpected, do expect DDR carriers to have higher TMB, but breast cancer has less TMB overall than other cancer types.

- Hypothesis: Somatic PIK3CA mutation carriers exhibit distinct gene expression patterns compared to PIK3CA wild‐type patients. Oncologist Comment: unexpected because would expect PIK3CA mutations would impact pathway gene expression. Would evlauate lobular vs ductal and HR+ vs HR- as this may be impacting differences.

- Hypothesis: ERBB2 (HER2) mRNA expression differs significantly across patients categorized by HER2 IHC status (Negative, Equivocal, Positive). Oncologist Comment: expected, IHC is protein mRNA can be evaried.

- Hypothesis: HER2 IHC score is positively correlated with ERBB2 (HER2) gene expression levels. Oncologist Comment: unexpected, would expect that HER2 IHC is associated with HER2 gene expression because amplification.

- Hypothesis: Breast cancer patients with high lymph node involvement (≥4 positive nodes) exhibit distinct gene expression signatures compared to those with low involvement (<4 positive nodes). Oncologist Comment: expected, pos axillary disease IS different from less axillary disease (would do 3 axillary nodes vs >3)

- Hypothesis: Patients with PIK3CA mutations present with lower pathologic T stage than PIK3CA wild‐type patients. Oncologist Comment: expect to have PIK3CA to have higher pathologic T stage, probably getting this numerical response bceause typically PIK3CA is checked in the metastatic stage in the community but the TCGA detected in earlier stages that would clinically be found

- Hypothesis: Breast tumors harboring PIK3CA mutations show altered expression of downstream PI3K-AKT pathway genes AKT1, MTOR, and RPS6KB1 compared to PIK3CA wild-type tumors. Oncologist Comment: expect this would be impacted, odd this is false.

- Hypothesis: Breast tumors harboring PIK3CA mutations exhibit higher expression of PIK3CA and its downstream effector AKT1 than wild‐type tumors. Oncologist Comment: would not call this false because AKT was abn expressed. Not unexpected.

- Hypothesis: Patients who received neoadjuvant treatment exhibit a different tumor mutational burden compared to those who did not receive neoadjuvant treatment. Oncologist Comment: Would not expect this. One modification would be TMB in patients receiving neoadjuvant chemotherapy between TNBC, HR+HER2-, HR+HER2+, and HR-HER2+

- Hypothesis: Tumor mutation burden (TMB) positively correlates with expression of the immune checkpoint gene PDCD1 (PD‐1) across TCGA‐BRCA patients. Oncologist Comment: Would evaluate this with the HR+ status, suspect would be more TNBC than HR+HER2-.

- Hypothesis: HER2-positive breast tumors exhibit lower CD8A expression (a marker of cytotoxic T cell infiltration) than HER2-negative tumors. Oncologist Comment: expected, there are some HER2+ Tumors with high TILS but overall would not expect to be too different, if compare HR+HER2 negative to HR+HER2+ to HR-HER2+ to TNBC would expect there be increasing CD8 T cells as loose HR (ER and PR) expression and go to TNBC.

- Hypothesis: Tumor infiltrating lymphocyte (TIL)–related gene signature expression associates with pathologic nodal stage (pathologic_n) independently of tumor size (pathologic_t). Oncologist Comment: Expected, TIL infiltration associated with good prognosis so would not expect nodal positivity with high TIL.

- Hypothesis: The gene expression profile of ER-positive breast tumors differs significantly from ER-negative tumors, with distinct sets of differentially expressed genes. Oncologist Comment: Expected and known. This is the definition of PAM50.= evaluation.

- Hypothesis: Mutation frequencies of PIK3CA and TP53 are significantly different between ER-positive and ER-negative breast tumors. Oncologist Comment: Expected, would not combine these two. Could look between lobular and ductal HR+HER2-.

- Hypothesis: Higher HER2 IHC score is associated with increased tumor ERBB2 (HER2) expression. Oncologist Comment: Would expect increased ERBB2 and IHC but not interesting question.

- Hypothesis: Estrogen receptor (ER) positive tumors exhibit higher tumor expression of the ESR1 gene than ER negative tumors. Oncologist Comment: No interesting question, the definition of ER- tumors is IHC expresison lacking protein expression of ER.

- Hypothesis: ERBB2 (HER2) mRNA expression levels are significantly higher in patients with HER2 IHC score 3+ or HER2 FISH positive status compared to those who are IHC 0/1+ and FISH negative. Oncologist Comment: expected, would be interesting to look at HER2 3+, HER2 2+ FISH pos, HER2 2+ FISH negative and HER2 1+ FISH negative particularly with Enhertu being effective in all of these.

##### Columns

| **Column Name** | **Description** |
| --- | --- |
| bcr_patient_barcode | TCGA patient barcode (BCR identifier) |
| gender | Reported gender |
| race | Race (list/label) |
| ethnicity | Ethnicity |
| days_to_birth | Negative days offset from birth |
| age_at_diagnosis | Age at initial pathologic diagnosis (years) |
| menopause_status | Menopause status at diagnosis |
| histological_type | Histological tumor type |
| diagnosis_year | Year of initial pathologic diagnosis |
| icd_10_code | ICD-10 disease classification code |
| tissue_source_site | Tissue source site code |
| prior_malignancy | History of prior malignancy diagnosis |
| er_status | Estrogen receptor status (Positive/Negative/Indeterminate) |
| pr_status | Progesterone receptor status (Positive/Negative/Indeterminate) |
| her2_ihc_status | HER2 immunohistochemistry receptor status |
| her2_fish_status | HER2 FISH (fluorescence in situ hybridization) result |
| her2_ihc_score | HER2 IHC score (0, 1+, 2+, 3+) |
| pathologic_stage | AJCC pathologic stage (post-surgery) |
| pathologic_t | AJCC pathologic T (tumor) stage |
| pathologic_n | AJCC pathologic N (lymph node) stage |
| pathologic_m | AJCC pathologic M (metastasis) stage |
| lymph_nodes_examined | Number of lymph nodes examined |
| lymph_nodes_positive | Number of lymph nodes positive by H&E |
| anatomic_subdivision | Anatomic neoplasm subdivision (breast quadrant/laterality) |
| tumor_status | Person neoplasm cancer status (tumor free/with tumor) |
| surgery_type | Breast carcinoma surgical procedure name |
| surgical_margin | Surgical margin status (Positive/Negative/Close) |
| neoadjuvant_treatment | History of neoadjuvant treatment |
| radiation_therapy | Radiation therapy administered (YES/NO) |
| pharmaceutical_therapy | Postoperative pharmaceutical therapy administered |
| drug_name | Primary drug name (first occurrence) |
| therapy_type | Primary therapy type (first occurrence) |
| vital_status | Patient vital status (Alive/Dead) |
| days_to_last_followup | Days from diagnosis to last follow-up |
| days_to_death | Days from diagnosis to death |
| new_tumor_event | New tumor event after initial treatment (YES/NO) |
| days_to_new_tumor | Days to new tumor event after initial treatment |

#### cleaned_gene_expression.csv

**dataset_info:**

name: TCGA-BRCA Gene Expression (RNA-Seq)

source: UCSC Xena - TOIL RSEM FPKM

normalization: RSEM expected_count, then log2(fpkm+0.001) transformed

platform: Illumina HiSeq RNA-Seq

data_type: Gene expression quantification

url: https://toil.xenahubs.net/download/tcga_RSEM_gene_fpkm.gz

**dimensions:**

genes: 60498

samples: 1068

description: Rows are genes (Ensembl gene IDs), columns are samples (TCGA patient

barcodes)

**sample_alignment:**

clinical_patients: 1068

matched_samples: 1068

note: Samples filtered to match patients in cleaned_clinical_data.csv

**data_description:**

values: Log2(FPKM + 0.001) - normalized gene expression levels

interpretation: Higher values indicate higher gene expression. Values are continuous

and log-transformed.

missing_data: Genes with all zeros or NaN across samples were removed

**usage_notes:**

- Gene identifiers are in the index (first column)

- Sample identifiers are in the column headers (TCGA patient barcodes)

- Data is pre-normalized and log-transformed, ready for analysis

- Match samples to clinical data using patient barcode (column headers)

- For patient-level analysis, data is already aggregated to one sample per patient

#### cleaned_mutations.csv

**dataset_info:**

name: TCGA-BRCA Somatic Mutations

source: GDC Data Portal - MC3 Public MAF

format: Mutation Annotation Format (MAF)

data_type: Somatic mutations (SNVs and small indels)

workflow: MC3 - Multi-Center Mutation Calling in Multiple Cancers

description: Curated somatic mutation calls combining multiple variant calling pipelines

**dimensions:**

total_mutations: 135030

unique_patients: 1020

unique_genes: 18907

columns: 21

**sample_alignment:**

clinical_patients: 1097

patients_with_mutations: 1020

note: Mutations filtered to match patients in cleaned_clinical_data.csv

**variant_summary:**

top_variant_classifications:

Missense_Mutation: 69328

Silent: 25786

3'UTR: 9585

Frame_Shift_Del: 9324

Nonsense_Mutation: 5890

Intron: 4617

5'UTR: 3484

Splice_Site: 2084

RNA: 1728

Frame_Shift_Ins: 961

3'Flank: 721

In_Frame_Del: 683

5'Flank: 606

Translation_Start_Site: 93

Nonstop_Mutation: 90

In_Frame_Ins: 50

variant_types:

SNP: 122135

DEL: 11564

INS: 1326

ONP: 5

description: Most common mutation types in the dataset

**top_mutated_genes:**

genes:

TTN: 446

PIK3CA: 425

TP53: 376

MUC16: 213

CDH1: 144

MAP3K1: 142

GATA3: 141

KMT2C: 132

RYR2: 121

SYNE1: 118

FLG: 107

DST: 106

HMCN1: 103

DMD: 92

NEB: 91

OBSCN: 91

USH2A: 88

RYR3: 85

SPTA1: 84

SYNE2: 80

description: Top 20 most frequently mutated genes

**column_descriptions:**

Hugo_Symbol: HUGO gene symbol

Chromosome: Chromosome where mutation occurs

Start_Position: Genomic start position (1-based)

Variant_Classification: Mutation type (e.g., Missense_Mutation, Nonsense_Mutation)

Variant_Type: SNP, DNP, INS, or DEL

HGVSp_Short: Protein change annotation

Tumor_Sample_Barcode: TCGA sample barcode

bcr_patient_barcode: Patient identifier (TCGA-XX-XXXX format)

**usage_notes:**

- Each row represents one somatic mutation

- Match to clinical data using the bcr_patient_barcode column

- For gene-level analysis, aggregate mutations by Hugo_Symbol

- For patient-level mutation burden, count mutations per bcr_patient_barcode

- Variant_Classification indicates functional impact

- FILTER column shows variant quality filtering status

##### Columns

| **Column Name** | **Description** |
| --- | --- |
| Hugo_Symbol | HUGO gene symbol |
| Chromosome | Chromosome where mutation occurs |
| Start_Position | Genomic start position (1-based coordinate) |
| End_Position | Genomic end position (1-based coordinate) |
| Strand | DNA strand on which the mutation is annotated (+ or -) |
| Variant_Classification | Functional class of the mutation (e.g., Missense_Mutation, Nonsense_Mutation, RNA, UTR, Frame_Shift) |
| Variant_Type | Type of variant call (e.g., SNP, DEL, INS, ONP) |
| Reference_Allele | Reference allele observed at the mutation site |
| Tumor_Seq_Allele1 | First observed tumor allele at the locus |
| Tumor_Seq_Allele2 | Second observed tumor allele at the locus |
| Tumor_Sample_Barcode | TCGA tumor sample barcode |
| bcr_patient_barcode | Patient identifier (TCGA-XX-XXXX format) |
| HGVSc | HGVS coding DNA change annotation (c.) using transcript coordinates |
| HGVSp | HGVS protein change annotation using full protein notation (p.) |
| HGVSp_Short | Shortened HGVS protein change notation |
| Transcript_ID | Ensembl transcript identifier (ENST...) used for annotation |
| Exon_Number | Exon where the variant occurs (e.g., 6/6) |
| IMPACT | Predicted functional impact class of the mutation (HIGH, MODERATE, LOW, MODIFIER) |
| SIFT | SIFT protein impact prediction in the form label(score), where label is one of tolerated/deleterious/deleterious_low_confidence and score is a probability (0–1) |
| PolyPhen | PolyPhen-2 protein impact prediction in the form label(score), where label is one of benign/possibly_damaging/probably_damaging and score is a probability (0–1) |
| FILTER | Variant filter status indicating whether the call passed QC filtering |

### Run Configuration (args.json)

| **Parameter** | **Value** |
| --- | --- |
| dataset_metadata | s3://ai2-asta-workspaces/autods/datasets/tcga-breast-cancer/tcga_breast_cancer_metadata_notes.json |
| out_dir | /results |
| model | o4-mini |
| belief_model | o4-mini |
| user_query | null |
| temperature | null |
| belief_temperature | null |
| reasoning_effort | medium |
| continue_from_dir | null |
| continue_from_json | null |
| n_experiments | 500 |
| k_experiments | 8 |
| allow_generate_experiments | True |
| n_belief_samples | 30 |
| timestamp_dir | True |
| exploration_weight | 2.0 |
| dataset_metadata_type | dbench |
| work_dir | work |
| delete_work_dir | True |
| mcts_selection | ucb1_recursive |
| pw_k | 1.0 |
| pw_alpha | 0.5 |
| beam_width | 8 |
| beam_branching_factor | null |
| k_parents | 3 |
| implicit_bayes_posterior | False |
| surprisal_width | 0.2 |
| belief_mode | boolean_cat |
| use_binary_reward | False |
| dedupe | True |
| only_save_results | False |
| experiment_first | False |
| code_timeout | 1800 |
| run_eda | False |
| n_warmstart | 20 |
| use_online_beliefs | False |
| evidence_weight | 2.0 |
| kl_scale | 5.0 |
| reward_mode | belief |
| warmstart_experiments | null |
| batch_size | 5 |
| silent | True |
| n_threads | 5 |
| async_prior | True |

**DataVoyager**

Here is the plan to follow as best as possible:

Objective: Determine whether histological subtype (ILC vs IDC) is associated with differential expression of checkpoint genes PDCD1 (PD-1) and CD274 (PD-L1) specifically in the HR+HER2- molecular subtype cohort.

Steps:

1. Filter data*1 to retain only samples classified as HR+HER2- (ER*STATUS = Positive, PR*STATUS = Positive, HER2*STATUS = Negative).
2. Within the filtered HR+HER2- cohort, identify and retain samples with histological subtype annotated as ILC or IDC.
3. Extract gene expression values for PDCD1 and CD274 from data_0 for the corresponding patient IDs identified in step 2.
4. Merge histological subtype (ILC vs IDC) with gene expression data (PDCD1 and CD274) for each sample.
5. Perform statistical analysis (e.g., t-test or non-parametric equivalent) to compare expression levels of PDCD1 and CD274 between ILC and IDC groups within HR+HER2- subtype.
6. Visualize results (e.g., boxplots or violin plots) to illustrate differences, if any, between ILC and IDC for both genes.
7. Summarize findings and interpret results, specifying whether significant differential expression exists between subtypes.

Deliverables:

1. Filtered and merged dataset containing HR+HER2- samples, histological subtype (ILC/IDC), and PDCD1/CD274 expression values.
2. Statistical test results comparing PDCD1 and CD274 expression between ILC and IDC within HR+HER2- subtype.
3. Visualizations (e.g., boxplots) of gene expression differences between subtypes.
4. Interpretation and summary addressing whether histological subtype is associated with differential expression of PDCD1 and/or CD274 among HR+HER2- samples.

Filter your clinical data to HR+HER2- cases (ER*STATUS == 'Positive', PR*STATUS == 'Positive', HER2*STATUS == 'Negative'), retaining only samples with HISTOLOGICAL*SUBTYPE of ILC or IDC. Extract matched PDCD1 and CD274 expression from the expression dataset, merge, then run suitable tests (e.g., Mann-Whitney U or t-test if justified) for expression differences between ILC and IDC for each gene. Summarize the result (group sizes, medians, p-values) and generate comparative boxplots.

**Function: exec_python**

**import** pandas **as** pd

**import** numpy **as** np

**import** matplotlib.pyplot **as** plt

**import** seaborn **as** sns

**from** scipy.stats **import** mannwhitneyu, ttest_ind, shapiro

\# 1. Filter clinical for HR+HER2- (strict: both ER and PR positive, HER2 negative)

hrp_her2n_mask = (

(data_1['ER_STATUS'] == 'Positive') &

(data_1['PR_STATUS'] == 'Positive') &

(data_1['HER2_STATUS'] == 'Negative')

)

clin_sub = data_1[hrp_her2n_mask].copy()

\# 2. Further filter for HISTOLOGICAL_SUBTYPE == 'Lobular' or 'Ductal/NST'

supported_subtypes = ['Lobular', 'Ductal/NST']

clin_sub = clin_sub[clin_sub['HISTOLOGICAL_SUBTYPE'].isin(supported_subtypes)]

\# 3. Extract/merge PDCD1 and CD274 expression

expr_df = data_0[data_0['Hugo_Symbol'].isin(['PDCD1', 'CD274'])]

expr_long = expr_df.set_index('Hugo_Symbol').T

expr_long.index.name = 'PATIENT_ID'

expr_long = expr_long.reset_index()

expr_long['PDCD1'] = pd.to_numeric(expr_long['PDCD1'], errors='coerce')

expr_long['CD274'] = pd.to_numeric(expr_long['CD274'], errors='coerce')

\# 4. Merge

merged = pd.merge(

clin_sub[['PATIENT_ID', 'HISTOLOGICAL_SUBTYPE']],

expr_long[['PATIENT_ID', 'PDCD1', 'CD274']],

on='PATIENT_ID', how='inner')

\# Prepare results dict for summaries

results = {}

plots = []

**for** gene **in** ['PDCD1', 'CD274']:

\# drop missing

subset = merged.dropna(subset=[gene, 'HISTOLOGICAL_SUBTYPE'])

group_ILC = subset[subset['HISTOLOGICAL_SUBTYPE'] == 'Lobular'][gene]

group_IDC = subset[subset['HISTOLOGICAL_SUBTYPE'] == 'Ductal/NST'][gene]

\# Check normality for each group

p_normal_ILC = shapiro(group_ILC)[1] **if** len(group_ILC) >= 3 **else** np.nan

p_normal_IDC = shapiro(group_IDC)[1] **if** len(group_IDC) >= 3 **else** np.nan

\# Mann-Whitney or t-test

**if** (p_normal_ILC > 0.05) **and** (p_normal_IDC > 0.05):

stat, pval = ttest_ind(group_ILC, group_IDC, nan_policy='omit')

test_type = 't-test'

**else**:

stat, pval = mannwhitneyu(group_ILC, group_IDC, alternative='two-sided')

test_type = 'Mann-Whitney U'

results[gene] = {

'n_ILC': len(group_ILC), 'median_ILC': np.median(group_ILC),

'n_IDC': len(group_IDC), 'median_IDC': np.median(group_IDC),

'test_type': test_type, 'statistic': stat, 'p_value': pval

}

\# boxplot

plt.figure(figsize=(7,5))

sns.boxplot(x='HISTOLOGICAL_SUBTYPE', y=gene, data=subset, order=['Ductal/NST', 'Lobular'])

plt.title(f'{gene} expression: HR+HER2- only')

plt.ylabel(f'{gene} expression')

plt.xlabel('Histological subtype')

plots.append(plt.show())

print('RESULTS (ILC vs IDC, HR+HER2- only):')

**for** gene, res **in** results.items():

print(f"{gene}: n_ILC={res['n_ILC']} median_ILC={res['median_ILC']:.3f} | n_IDC={res['n_IDC']} median_IDC={res['median_IDC']:.3f} | {res['test_type']} statistic={res['statistic']:.3f} p={res['p_value']:.4g}")

**all ID: call_NKhswbtL4QBr7pj2T7U1ezX3**

Execution successful.

Output:

RESULTS (ILC vs IDC, HR+HER2- only):

PDCD1: n_ILC=74 median_ILC=5.540 | n_IDC=696 median_IDC=5.482 | Mann-Whitney U statistic=29608.000 p=0.03405

CD274: n_ILC=74 median_ILC=5.509 | n_IDC=696 median_IDC=5.450 | Mann-Whitney U statistic=29664.000 p=0.03154

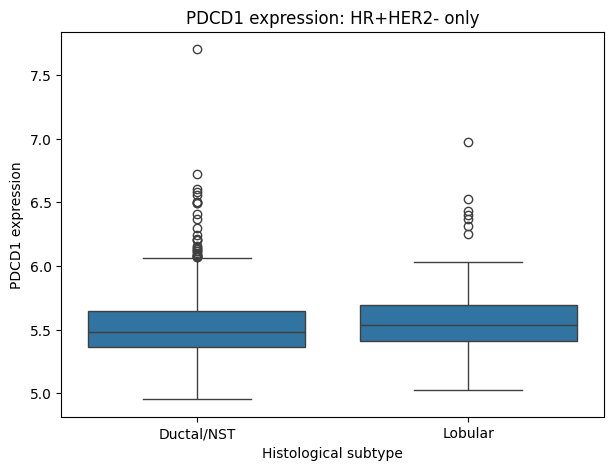

**Interpretation for Plot 1:**

1. Plot Type: The plot is a box plot, used for comparing the distribution of PDCD1 expression between two histological subtypes (Ductal/NST and Lobular) within HR+HER2- only cases.
2. Axes:
   - X-axis: Labeled 'Histological subtype' with categories 'Ductal/NST' and 'Lobular'.
   - Y-axis: Labeled 'PDCD1 expression', numeric range from approximately 4.8 to 7.8 (no units specified).
3. Data Trends:
   - Both groups have similar interquartile ranges and median values.
   - The Ductal/NST group shows more outliers above the upper whisker compared to the Lobular group.
   - Both groups' distributions appear slightly right-skewed as indicated by the presence of outliers above the boxes.
4. Annotations and Legends: No specific annotations or legends are present, but the central horizontal line inside each box indicates the median.
5. Overall Impression: PDCD1 expression shows comparable distributions between histological subtypes, with Ductal/NST exhibiting slightly more high-value outliers.

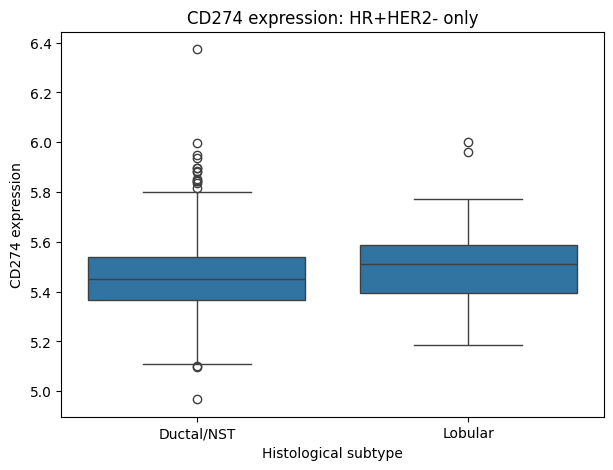

**Interpretation for Plot 2:**

1. Plot Type: The plot is a grouped boxplot, showing the distribution of CD274 expression across two histological subtypes for the HR+HER2- only group. Boxplots are typically used to visualize the central tendency, variability, and potential outliers in a continuous variable for different categories.
2. Axes:
   - X-axis: Labeled 'Histological subtype' with categories 'Ductal/NST' and 'Lobular'. No units are necessary as these are categorical.
   - Y-axis: Labeled 'CD274 expression' (no units provided), with a value range from approximately 5.0 to 6.4.
3. Data Trends: Both subtypes have similar medians around 5.5–5.6. Ductal/NST shows more outliers above the upper whisker (a cluster between ~5.85 & 6) and a few lower outliers close to 5.0. Lobular shows slightly fewer outliers but a similar interquartile range. The overall spread and median are quite comparable.
4. Annotations and Legends: There is no legend, but the title 'CD274 expression: HR+HER2- only' serves as annotation/context. Outliers are marked as individual dots.
5. Overall Impression: There are minimal differences in CD274 expression distributions between Ductal/NST and Lobular histological subtypes within the HR+HER2- group; expression levels and variance appear broadly comparable, warranting further statistical analysis for subtle differences.
6. HR+HER2- molecular subtype is defined as ERSTATUS and PRSTATUS both 'Positive' and HER2_STATUS 'Negative'.
7. Histological subtype is accurately captured as 'Lobular' (ILC) and 'Ductal/NST' (IDC) in the clinical data.
8. PDCD1 and CD274 gene expression values are normalized and directly comparable across samples.
9. Sample sizes for ILC (n=74) and IDC (n=696) within HR+HER2- are sufficient for Mann-Whitney U testing.
10. Nonparametric tests are appropriate due to non-normality of expression distributions.
11. Only primary tumor samples are included and are comparable.
12. No major batch effects or confounders are present beyond those controlled for.
13. Yes, within the HR+HER2- molecular subtype in the METABRIC dataset, ILC (Invasive Lobular Carcinoma) shows a small but statistically significant increase in expression of both PDCD1 (PD-1) and CD274 (PD-L1) compared to IDC (Invasive Ductal Carcinoma). Median PDCD1 expression is 5.54 in ILC vs 5.48 in IDC (p=0.034), and median CD274 is 5.51 in ILC vs 5.45 in IDC (p=0.032) by Mann-Whitney U test. However, the difference is small and distributions are highly overlapping, so the biological significance is uncertain.
14. **Generated code**
15. This is an aggregation of all of the code snippets output in the Reasoning section
16. **import** pandas **as** pd
18. # Preview columns in clinical dataset to identify appropriate columns
19. print('Columns in data_1:')
20. print(data_1.columns.tolist())
22. # Get all unique values from each column in data_1 to help confirm names of relevant columns
23. print('\nUnique values per column:')
24. **for** col **in** data_1.columns:
25. print(col, ':', data_1[col].unique()[:5]) # show up to 5 unique values for brevity
27. # Identify gene rows for PDCD1 and CD274 in data_0
28. print('\nUnique Hugo_Symbols in data_0:')
29. print(data_0['Hugo_Symbol'].unique()[:10]) # Show first 10 unique gene symbols
31. genes_of_interest = ['PDCD1', 'CD274']
32. gene_mask = data_0['Hugo_Symbol'].isin(genes_of_interest)
33. data_genes = data_0[gene_mask]
35. print('\nGenes of interest found:')
36. print(data_genes['Hugo_Symbol'].tolist())
38. # Reshape gene data: index = gene, columns = sample IDs
39. data_genes_long = data_genes.set_index('Hugo_Symbol').T
40. # Remove possible index name for clarity
41. data_genes_long.index.name = **None**
43. print('\nGene expression DataFrame shape:', data_genes_long.shape)
44. print('First few rows of reshaped gene data:')
45. print(data_genes_long.head())
47. # Merge expression data (index = sample ID) with clinical data (PATIENT_ID)
48. data_genes_long = data_genes_long.rename_axis('PATIENT_ID').reset_index()
49. merged = pd.merge(data_1, data_genes_long, on='PATIENT_ID', how='inner')
51. # Guess columns for required variables
52. desired_cols = ['PATIENT_ID', 'HISTOLOGICAL_SUBTYPE', 'CLAUDIN_SUBTYPE', 'PDCD1', 'CD274']
53. existing_cols = [col **for** col **in** desired_cols **if** col **in** merged.columns]
54. print('\nColumns present in merged dataset:', existing_cols)
56. # Subset merged to only non-missing for all analysis variables
57. analysis_vars = existing_cols
58. analysis_df = merged.dropna(subset=analysis_vars)
60. print('\nShape of cleaned analysis DataFrame:', analysis_df.shape)
61. print('First few rows of analysis DataFrame:')
62. print(analysis_df[analysis_vars].head())
64. **import** pandas **as** pd
65. **import** matplotlib.pyplot **as** plt
66. **import** seaborn **as** sns
67. **import** scipy.stats **as** stats
68. **import** statsmodels.formula.api **as** smf
70. # For concise output
71. pd.set_option('display.max_columns', 15)
73. # --- Use data from previous step (analysis_df) ---
75. # 1. Group size/summary tables
76. summary = analysis_df.groupby(['HISTOLOGICAL_SUBTYPE', 'CLAUDIN_SUBTYPE']).agg(
77. count = ('PATIENT_ID', 'count'),
78. pdcd1_mean = ('PDCD1', 'mean'),
79. pdcd1_std = ('PDCD1', 'std'),
80. cd274_mean = ('CD274', 'mean'),
81. cd274_std = ('CD274', 'std')
82. ).reset_index()
84. print('Summary table by histological and molecular subtype:')
85. print(summary)
87. # 2. Plot stratified distributions for PDCD1 and CD274
88. fig, axs = plt.subplots(1, 2, figsize=(16,6))
90. sns.boxplot(
91. x='HISTOLOGICAL_SUBTYPE', y='PDCD1', hue='CLAUDIN_SUBTYPE',
92. data=analysis_df, ax=axs[0])
93. axs[0].set_title('PDCD1 Expression by Histological and Molecular Subtype')
94. axs[0].set_ylabel('PDCD1 Expression')
95. axs[0].tick_params(axis='x', rotation=30)
97. sns.boxplot(
98. x='HISTOLOGICAL_SUBTYPE', y='CD274', hue='CLAUDIN_SUBTYPE',
99. data=analysis_df, ax=axs[1])
100. axs[1].set_title('CD274 Expression by Histological and Molecular Subtype')
101. axs[1].set_ylabel('CD274 Expression')
102. axs[1].tick_params(axis='x', rotation=30)
104. plt.tight_layout()
105. _stratified_plot = plt.show()
107. # 3. Distribution checks (normality, variance) - PDCD1 and CD274
108. print('\nNormality test results (Shapiro-Wilk, all data):')
109. **for** gene **in** ['PDCD1', 'CD274']:
110. w, pval = stats.shapiro(analysis_df[gene])
111. print(f'{gene}: W={w:.3f}, p={pval:.4f}')
112. print('\nVariance summary:')
113. **for** gene **in** ['PDCD1', 'CD274']:
114. print(f"{gene} variance: {analysis_df[gene].var():.3f}")
116. # 4. Linear modeling (GLM, controlling for molecular subtype)
117. print('\nLinear regression results controlling for CLAUDIN_SUBTYPE:')
118. **for** gene **in** ['PDCD1', 'CD274']:
119. print(f'\n--- {gene} ---')
120. formula = f'{gene} ~ C(HISTOLOGICAL_SUBTYPE) + C(CLAUDIN_SUBTYPE)'
121. model = smf.ols(formula=formula, data=analysis_df).fit()
122. print(model.summary())
124. # Extract effect sizes and p-values for HISTOLOGICAL_SUBTYPE
125. histo_effects = model.params.filter(like='HISTOLOGICAL_SUBTYPE')
126. histo_pvals = model.pvalues.filter(like='HISTOLOGICAL_SUBTYPE')
127. **for** k **in** histo_effects.index:
128. print(f'Effect size for {k}: {histo_effects[k]:.3f}, p-value: {histo_pvals[k]:.4f}')
130. **import** pandas **as** pd
131. # 1. Identify columns for expression extraction and clinical column names
132. print('data_1 columns:', data_1.columns.tolist())
133. print('First few unique HISTOLOGICAL_SUBTYPE:', data_1['HISTOLOGICAL_SUBTYPE'].unique())
134. print('Unique values for ER_STATUS:', data_1['ER_STATUS'].unique())
135. print('Unique values for PR_STATUS:', data_1['PR_STATUS'].unique())
136. print('Unique values for HER2_STATUS:', data_1['HER2_STATUS'].unique())
138. # Attempt to find if there is a column with exactly the desired strings
139. **if** 'HISTOLOGICAL_SUBTYPE' **in** data_1.columns:
140. hist_col = 'HISTOLOGICAL_SUBTYPE'
141. **else**:
142. possible_cols = [col **for** col **in** data_1.columns **if** 'hist' **in** col.lower()]
143. print('Possible histo col:', possible_cols)
144. hist_col = possible_cols[0] **if** possible_cols **else** **None**
146. # 2. Subset to only lobular or ductal carcinoma (allowing for possible synonyms)
147. hist_map = {
148. 'Infiltrating lobular carcinoma': ['Lobular', 'Infiltrating lobular carcinoma'],
149. 'Infiltrating ductal carcinoma': ['Ductal/NST', 'Infiltrating ductal carcinoma']
150. }
152. mask = data_1[hist_col].isin(hist_map['Infiltrating lobular carcinoma'] + hist_map['Infiltrating ductal carcinoma'])
153. data1_sub = data_1.loc[mask].copy()
155. # 3. Molecular subtype variable: HR = ER or PR positive
157. **def** **molecular_subtype**(row):
158. hrp = (str(row['ER_STATUS']).strip().lower() == 'positive') **or** (str(row['PR_STATUS']).strip().lower() == 'positive')
159. her2p = (str(row['HER2_STATUS']).strip().lower() == 'positive')
160. **if** hrp **and** **not** her2p:
161. **return** 'HR+HER2-'
162. **elif** hrp **and** her2p:
163. **return** 'HR+HER2+'
164. **elif** (**not** hrp) **and** her2p:
165. **return** 'HR-HER2+'
166. **elif** (**not** hrp) **and** (**not** her2p):
167. **return** 'TNBC'
168. **else**:
169. **return** 'Unknown'
171. # Apply molecular subtype logic
172. data1_sub['molecular_subtype'] = data1_sub.apply(molecular_subtype, axis=1)
174. print('Molecular subtype crosstab:')
175. print(pd.crosstab(data1_sub[hist_col], data1_sub['molecular_subtype']))
177. # 4. Get PDCD1 and CD274 gene rows (by Ensembl ID in data_0)
178. ensembl_ids = ['ENSG00000188389', 'ENSG00000120217']
179. **if** 'Hugo_Symbol' **in** data_0.columns:
180. gene_mask = data_0['Hugo_Symbol'].isin(ensembl_ids)
181. data0_genes = data_0[gene_mask]
182. gene_labels = dict(zip(ensembl_ids, ['PDCD1_expression', 'CD274_expression']))
183. data0_genes = data0_genes.assign(Gene=data0_genes['Hugo_Symbol'].map(gene_labels))
184. **else**:
185. print('Gene identifier column not present')
186. data0_genes = pd.DataFrame([])
188. # Transpose so index is sample (patient) id, columns are expression for each gene
189. data0_long = data0_genes.set_index('Gene').drop('Hugo_Symbol', axis=1).T
190. # Remove index name for clarity
191. data0_long.index.name = **None**
193. # 5. Merge gene expression (per patient) with clinical
194. data0_long = data0_long.rename_axis('PATIENT_ID').reset_index()
195. merged = pd.merge(data1_sub, data0_long, on='PATIENT_ID', how='inner')
197. sel_cols = ['PATIENT_ID', hist_col, 'AGE_AT_DIAGNOSIS', 'molecular_subtype', 'PDCD1_expression', 'CD274_expression']
198. output_cols = [col **for** col **in** sel_cols **if** col **in** merged.columns]
199. merged_clean = merged[output_cols].dropna()
201. print('Shape of final cleaned/merged dataframe:', merged_clean.shape)
202. print('First few rows:')
203. print(merged_clean.head())
205. **import** pandas **as** pd
206. **import** seaborn **as** sns
207. **import** matplotlib.pyplot **as** plt
208. **import** statsmodels.formula.api **as** smf
209. **import** scipy.stats **as** stats
211. # We'll continue from merged_clean (already has required columns/expressions)
212. print('Merged/cleaned dataframe columns:', merged_clean.columns.tolist())
214. # Rename expression columns for clarity (repeat extraction in case columns weren't included before)
215. # Redo extraction in case not present
216. gene_map = {'ENSG00000188389': 'PDCD1_expression', 'ENSG00000120217': 'CD274_expression'}
217. **if** 'PDCD1_expression' **not** **in** merged_clean.columns **or** 'CD274_expression' **not** **in** merged_clean.columns:
218. ids_to_genes = {'ENSG00000188389': 'PDCD1', 'ENSG00000120217': 'CD274'}
219. gene_rows = data_0[data_0['Hugo_Symbol'].isin(ids_to_genes.keys())]
220. gene_rows = gene_rows.assign(Gene=gene_rows['Hugo_Symbol'].map(ids_to_genes))
221. expr_df = gene_rows.set_index('Gene').drop('Hugo_Symbol', axis=1).T
222. expr_df.index.name = 'PATIENT_ID'
223. expr_df = expr_df.reset_index()
224. merged_clean = pd.merge(merged_clean, expr_df, on='PATIENT_ID', how='left')
225. merged_clean = merged_clean.rename(columns={'PDCD1': 'PDCD1_expression', 'CD274': 'CD274_expression'})
227. # Remove rows with missing expression values
228. merged_clean2 = merged_clean.dropna(subset=['PDCD1_expression', 'CD274_expression'])
230. # Regression for each gene
231. regression_results = {}
232. **for** gene **in** ['PDCD1_expression', 'CD274_expression']:
233. print(f'\n==== Linear model for {gene} ====')
234. formula = f'{gene} ~ C(HISTOLOGICAL_SUBTYPE) + C(molecular_subtype) + AGE_AT_DIAGNOSIS'
235. model = smf.ols(formula, data=merged_clean2).fit()
236. regression_results[gene] = model
237. print(model.summary())
238. # Residual plots: normality and homoscedasticity
239. residuals = model.resid
240. fitted = model.fittedvalues
241. fig, axs = plt.subplots(1, 2, figsize=(12,5))
242. sns.histplot(residuals, kde=**True**, ax=axs[0])
243. axs[0].set_title(f'Residual Histogram ({gene})')
244. stats.probplot(residuals, dist="norm", plot=axs[1])
245. axs[1].set_title(f'QQ-plot Residuals ({gene})')
246. plt.tight_layout()
247. _ = plt.show()
248. fig, ax = plt.subplots(figsize=(6,5))
249. ax.scatter(fitted, residuals)
250. ax.set_xlabel('Fitted values')
251. ax.set_ylabel('Residuals')
252. ax.set_title(f'Residuals vs Fitted ({gene})')
253. _ = plt.show()
254. # Additional test for normality
255. shapiro = stats.shapiro(residuals)
256. print(f'{gene} residual Shapiro-Wilk: W={shapiro[0]:.3f}, p-value={shapiro[1]:.5f}')
258. # Wilcoxon rank-sum if any violate normality (p < 0.05)
259. print('\n== Wilcoxon tests by molecular subtype (if needed) ==')
260. **for** gene **in** ['PDCD1_expression', 'CD274_expression']:
261. res = regression_results[gene]
262. residuals = res.resid
263. shapiro_p = stats.shapiro(residuals)[1]
264. **if** shapiro_p < 0.05:
265. **for** subtype **in** merged_clean2['molecular_subtype'].unique():
266. sub = merged_clean2[merged_clean2['molecular_subtype'] == subtype]
267. groups = sub['HISTOLOGICAL_SUBTYPE'].unique()
268. **if** len(groups) == 2:
269. group1 = sub[sub['HISTOLOGICAL_SUBTYPE'] == groups[0]][gene].dropna()
270. group2 = sub[sub['HISTOLOGICAL_SUBTYPE'] == groups[1]][gene].dropna()
271. **if** len(group1) > 2 **and** len(group2) > 2:
272. stat, p = stats.ranksums(group1, group2)
273. print(f'Wilcoxon rank-sum for {gene}, subtype {subtype}, {groups[0]} vs {groups[1]}: statistic={stat:.3f}, p-value={p:.4f}')
274. **else**:
275. print(f'Not enough samples for {gene}, subtype {subtype}')
277. **import** pandas **as** pd
278. # 1. Extract PDCD1 and CD274 (by Ensembl ID)
279. ensembl_ids = ['ENSG00000188389', 'ENSG00000120217']
280. expr_rows = data_0[data_0['Hugo_Symbol'].isin(ensembl_ids)]
281. expr_map = {'ENSG00000188389': 'PDCD1_expression', 'ENSG00000120217': 'CD274_expression'}
282. expr_rows = expr_rows.assign(Gene=expr_rows['Hugo_Symbol'].map(expr_map))
284. # 2. Transpose so index=sample (e.g., MB-0000), columns=['PDCD1_expression', 'CD274_expression']
285. expr_t = expr_rows.set_index('Gene').drop('Hugo_Symbol', axis=1).T
286. expr_t.index.name = 'PATIENT_ID'
287. expr_t = expr_t.reset_index()
289. # 3. Merge with cleaned clinical dataframe (merged_clean)
290. merged_with_expr = pd.merge(merged_clean, expr_t, on='PATIENT_ID', how='inner')
291. print('Column names:', merged_with_expr.columns.tolist())
292. print('First few rows:')
293. print(merged_with_expr.head())
295. # Extract gene expression rows for PDCD1 and CD274
296. ensembl_ids = ['ENSG00000188389', 'ENSG00000120217']
297. gene_labels = {'ENSG00000188389': 'PDCD1_expression', 'ENSG00000120217': 'CD274_expression'}
298. expr_rows = data_0[data_0['Hugo_Symbol'].isin(ensembl_ids)]
299. expr_rows = expr_rows.assign(Gene=expr_rows['Hugo_Symbol'].map(gene_labels))
301. # Transpose to have patient IDs as rows and genes as columns
302. expr_t = expr_rows.set_index('Gene').drop('Hugo_Symbol', axis=1).T
303. expr_t.index.name = 'PATIENT_ID'
304. expr_t = expr_t.reset_index()
306. # Merge with cleaned/merged clinical dataframe on PATIENT_ID
307. merged_final = pd.merge(merged_clean, expr_t, on='PATIENT_ID', how='inner')
308. print('Final merged column names:', merged_final.columns.tolist())
309. print('First five rows:')
310. print(merged_final.head())
312. # Extract gene expression rows for PDCD1 and CD274 (by Ensembl ID)
313. ensembl_ids = ['ENSG00000188389', 'ENSG00000120217']
314. gene_labels = {'ENSG00000188389': 'PDCD1_expression', 'ENSG00000120217': 'CD274_expression'}
315. expr_rows = data_0[data_0['Hugo_Symbol'].isin(ensembl_ids)]
316. expr_rows = expr_rows.assign(Gene=expr_rows['Hugo_Symbol'].map(gene_labels))
318. # Transpose with patient IDs as rows and genes as columns
319. expr_t = expr_rows.set_index('Gene').drop('Hugo_Symbol', axis=1).T
320. expr_t.index.name = 'PATIENT_ID'
321. expr_t = expr_t.reset_index()
323. # Merge with merged_clean (clinical subset)
324. merged_final = pd.merge(merged_clean, expr_t, on='PATIENT_ID', how='inner')
326. # Drop any rows with missing gene expression values
327. merged_final = merged_final.dropna(subset=['PDCD1_expression', 'CD274_expression'])
329. print('Columns present in merged_final:', merged_final.columns.tolist())
330. print('First five rows:')
331. print(merged_final.head())
333. # Step 1: Extract the rows for PDCD1 and CD274 by Ensembl ID
334. ens_pdcd1 = 'ENSG00000188389'
335. ens_cd274 = 'ENSG00000120217'
336. expr_pdcd1 = data_0[data_0['Hugo_Symbol'] == ens_pdcd1].iloc[:, 1:].T
337. expr_cd274 = data_0[data_0['Hugo_Symbol'] == ens_cd274].iloc[:, 1:].T
339. # Step 2: Set columns and index for merging
340. expr_pdcd1.columns = ['PDCD1_expression']
341. expr_pdcd1.index.name = 'PATIENT_ID'
342. expr_cd274.columns = ['CD274_expression']
343. expr_cd274.index.name = 'PATIENT_ID'
345. # Step 3: Merge the two gene dataframes (on PATIENT_ID)
346. gene_expr = pd.merge(expr_pdcd1, expr_cd274, left_index=**True**, right_index=**True**)
347. gene_expr = gene_expr.reset_index()
349. # Step 4: Merge with clinical and drop rows with missing gene expression values
350. final = pd.merge(merged_clean, gene_expr, on='PATIENT_ID', how='inner')
351. final = final.dropna(subset=['PDCD1_expression', 'CD274_expression'])
353. print('Merged dataframe columns:', final.columns.tolist())
354. print('First five rows:')
355. print(final.head())
357. # 1. Print unique 'Hugo_Symbol' values to verify gene naming
358. unique_symbols = data_0['Hugo_Symbol'].unique()
359. print('First 20 unique Hugo_Symbol values:', unique_symbols[:20])
361. # Check if PDCD1 or CD274 or similar symbols are in Hugo_Symbol
362. print("Contains 'PDCD1'?", 'PDCD1' **in** unique_symbols)
363. print("Contains 'CD274'?", 'CD274' **in** unique_symbols)
365. # Stop here and examine output before attempting to extract rows
367. # Extract gene symbol expression for PDCD1 and CD274
368. expr_rows = data_0[data_0['Hugo_Symbol'].isin(['PDCD1', 'CD274'])]
369. expr_rows = expr_rows.set_index('Hugo_Symbol')
370. expr_rows_t = expr_rows.transpose()
371. # First row is now 'PDCD1', second is 'CD274', rest are patient IDs
372. expr_rows_t = expr_rows_t.iloc[1:, :] # skip the index row
373. expr_rows_t.index.name = 'PATIENT_ID'
374. expr_rows_t = expr_rows_t.rename(columns={'PDCD1': 'PDCD1_expression', 'CD274': 'CD274_expression'})
375. expr_rows_t = expr_rows_t.reset_index()
376. # Convert expression columns to float
377. **for** col **in** ['PDCD1_expression', 'CD274_expression']:
378. expr_rows_t[col] = pd.to_numeric(expr_rows_t[col], errors='coerce')
380. # Merge with clinical subset on PATIENT_ID
381. merged_with_expr = pd.merge(merged_clean, expr_rows_t, on='PATIENT_ID', how='inner')
383. print('Final merged dataframe columns:', merged_with_expr.columns.tolist())
384. print('First five rows:')
385. print(merged_with_expr.head())
386. print('Any missing values in PDCD1_expression:', merged_with_expr['PDCD1_expression'].isna().any())
387. print('Any missing values in CD274_expression:', merged_with_expr['CD274_expression'].isna().any())
389. **import** statsmodels.formula.api **as** smf
390. **import** matplotlib.pyplot **as** plt
391. **import** seaborn **as** sns
392. **import** scipy.stats **as** stats
393. **import** pandas **as** pd
395. # Data is in merged_with_expr, use only complete cases for analysis
396. model_df = merged_with_expr.dropna(subset=['PDCD1_expression', 'CD274_expression',
397. 'HISTOLOGICAL_SUBTYPE', 'molecular_subtype', 'AGE_AT_DIAGNOSIS'])
398. # Linear regression for PDCD1_expression
399. glm_pdcd1 = smf.ols('PDCD1_expression ~ C(HISTOLOGICAL_SUBTYPE) + C(molecular_subtype) + AGE_AT_DIAGNOSIS', data=model_df).fit()
400. print('\n=== Linear Model: PDCD1_expression ===')
401. print(glm_pdcd1.summary())
403. # Linear regression for CD274_expression
404. glm_cd274 = smf.ols('CD274_expression ~ C(HISTOLOGICAL_SUBTYPE) + C(molecular_subtype) + AGE_AT_DIAGNOSIS', data=model_df).fit()
405. print('\n=== Linear Model: CD274_expression ===')
406. print(glm_cd274.summary())
408. # Diagnostics: residual normality
409. **for** gene, model **in** [('PDCD1_expression', glm_pdcd1), ('CD274_expression', glm_cd274)]:
410. residuals = model.resid
411. fitted = model.fittedvalues
412. print(f"\nResidual Shapiro-Wilk for {gene}:")
413. shapiro = stats.shapiro(residuals)
414. print(f"W={shapiro.statistic:.4f}, p={shapiro.pvalue:.4e}")
415. # Plot residual histogram and Q-Q plot
416. fig, axs = plt.subplots(1, 2, figsize=(12,5))
417. sns.histplot(residuals, kde=**True**, ax=axs[0])
418. axs[0].set_title(f'{gene} Residuals Histogram')
419. stats.probplot(residuals, dist="norm", plot=axs[1])
420. axs[1].set_title(f'{gene} Residuals QQ-Plot')
421. plt.tight_layout()
422. _ = plt.show()
423. # Plot residuals vs fitted
424. fig, ax = plt.subplots(figsize=(6,5))
425. ax.scatter(fitted, residuals, alpha=0.7)
426. ax.axhline(0, color='grey', linestyle='--')
427. ax.set_xlabel('Fitted Values')
428. ax.set_ylabel('Residuals')
429. ax.set_title(f'{gene} Residuals vs Fitted')
430. plt.tight_layout()
431. _ = plt.show()
433. # If residuals violate normality (p < 0.05), do Wilcoxon tests within each molecular subtype
434. # Only perform if either Shapiro p<0.05
435. do_pdcd1 = stats.shapiro(glm_pdcd1.resid).pvalue < 0.05
436. do_cd274 = stats.shapiro(glm_cd274.resid).pvalue < 0.05
437. **if** do_pdcd1 **or** do_cd274:
438. print("\nStratified Wilcoxon rank-sum test results:")
439. **for** gene **in** ['PDCD1_expression', 'CD274_expression']:
440. **if** (gene == 'PDCD1_expression' **and** do_pdcd1) **or** (gene == 'CD274_expression' **and** do_cd274):
441. **for** subtype **in** sorted(model_df['molecular_subtype'].unique()):
442. sub = model_df[model_df['molecular_subtype'] == subtype]
443. types = sub['HISTOLOGICAL_SUBTYPE'].unique()
444. **if** len(types) == 2:
445. vals1 = sub[sub['HISTOLOGICAL_SUBTYPE'] == types[0]][gene]
446. vals2 = sub[sub['HISTOLOGICAL_SUBTYPE'] == types[1]][gene]
447. **if** len(vals1) >= 3 **and** len(vals2) >= 3:
448. stat, p = stats.ranksums(vals1, vals2)
449. print(f"{gene}, {subtype}: {types[0]} vs {types[1]} Wilcoxon statistic={stat:.3f}, p-value={p:.4e}")
450. **else**:
451. print(f"{gene}, {subtype}: Not enough samples for Wilcoxon test")
453. # Interpretation summary
454. **def** **interpretation**():
455. print('\nINTERPRETATION:')
456. print('Each gene (PDCD1, CD274) was regressed on histological subtype, molecular subtype, and age at diagnosis.\n')
457. print('Examine p-values and effect estimates for the histological subtype variable. If p > 0.05, histological subtype is not significantly associated with gene expression;\nif p < 0.05, there is a significant difference after adjustment. Non-normality indicates that Wilcoxon results are also relevant.\n')
458. print('The molecular subtype covariate should be expected to have a strong influence (esp. high for TNBC/HR- and/or claudin-low if included).')
459. print('Model diagnostics and Wilcoxon (if run) indicate how robust these findings are to violations of modeling assumptions.')
460. interpretation()
462. **import** pandas **as** pd
463. **import** numpy **as** np
464. **import** matplotlib.pyplot **as** plt
465. **import** seaborn **as** sns
466. **from** scipy.stats **import** mannwhitneyu, ttest_ind
468. # Define clinical groups in data_1
469. clin = data_1.copy()
470. # TNBC
471. mask_tnbc = (
472. (clin['ER_STATUS'] == 'Negative') &
473. (clin['PR_STATUS'] == 'Negative') &
474. (clin['HER2_STATUS'] == 'Negative')
475. )
476. # HR+HER2–
477. mask_hrp_her2n = (
478. ((clin['ER_STATUS'] == 'Positive') | (clin['PR_STATUS'] == 'Positive')) &
479. (clin['HER2_STATUS'] == 'Negative')
480. )
481. # List of sample IDs for each group
482. tnbc_ids = clin.loc[mask_tnbc, 'PATIENT_ID'].values
483. hrp_her2n_ids = clin.loc[mask_hrp_her2n, 'PATIENT_ID'].values
485. # 1. Extract PDCD1 row from data_0
486. pdcd1_row = data_0[data_0['Hugo_Symbol'] == 'PDCD1']
487. expr = pdcd1_row.drop('Hugo_Symbol', axis=1).T # rows: sample IDs
488. expr.index.name = 'PATIENT_ID'
489. expr.columns = ['PDCD1_expression']
490. expr = expr.reset_index()
491. expr['PDCD1_expression'] = pd.to_numeric(expr['PDCD1_expression'], errors='coerce')
493. # 2. Map expression to groups
494. expr_tnbc = expr[expr['PATIENT_ID'].isin(tnbc_ids)].dropna(subset=['PDCD1_expression'])
495. expr_hrp_her2n = expr[expr['PATIENT_ID'].isin(hrp_her2n_ids)].dropna(subset=['PDCD1_expression'])
497. # 3. Group sizes
498. n_tnbc = expr_tnbc.shape[0]
499. n_hrp_her2n = expr_hrp_her2n.shape[0]
501. # 4. Stats
502. **def** **describe**(x):
503. **return** {
504. 'n': len(x),
505. 'median': np.median(x),
506. 'mean': np.mean(x),
507. 'IQR': np.percentile(x, 75) - np.percentile(x, 25)
508. }
509. desc_tnbc = describe(expr_tnbc['PDCD1_expression'])
510. desc_hrp_her2n = describe(expr_hrp_her2n['PDCD1_expression'])
512. print('Group sizes - TNBC:', n_tnbc, 'HR+HER2-', n_hrp_her2n)
513. print('PDCD1 stats for TNBC:', desc_tnbc)
514. print('PDCD1 stats for HR+HER2-:', desc_hrp_her2n)
516. # 5. Boxplot
517. plt.figure(figsize=(8,6))
518. plot_data = pd.concat([
519. pd.DataFrame({'group':'TNBC','expr':expr_tnbc['PDCD1_expression']}),
520. pd.DataFrame({'group':'HR+HER2-','expr':expr_hrp_her2n['PDCD1_expression']})
521. ])
522. sns.boxplot(x='group', y='expr', data=plot_data)
523. plt.title('PDCD1 expression in TNBC vs HR+HER2-')
524. plt.ylabel('PDCD1 Expression')
525. _ = plt.show()
527. # 6. Statistical test: Normality check
528. **from** scipy.stats **import** shapiro
529. stat_t, pval_t = shapiro(expr_tnbc['PDCD1_expression'])
530. stat_h, pval_h = shapiro(expr_hrp_her2n['PDCD1_expression'])
532. print(f'Shapiro normality p (TNBC): {pval_t:.4g}, HR+HER2-: {pval_h:.4g}')
533. # If either group non-normal, use Mann-Whitney
534. **if** pval_t < 0.05 **or** pval_h < 0.05:
535. stat, pval = mannwhitneyu(expr_tnbc['PDCD1_expression'], expr_hrp_her2n['PDCD1_expression'], alternative='two-sided')
536. print(f"Mann-Whitney U test: statistic={stat:.3f}, p-value={pval:.4e}")
537. **else**:
538. stat, pval = ttest_ind(expr_tnbc['PDCD1_expression'], expr_hrp_her2n['PDCD1_expression'])
539. print(f"t-test: statistic={stat:.3f}, p-value={pval:.4e}")
541. **import** pandas **as** pd
542. **import** numpy **as** np
543. **import** matplotlib.pyplot **as** plt
544. **import** seaborn **as** sns
545. **from** scipy.stats **import** mannwhitneyu, ttest_ind, shapiro
547. # 1. Filter clinical for HR+HER2- (strict: both ER and PR positive, HER2 negative)
548. hrp_her2n_mask = (
549. (data_1['ER_STATUS'] == 'Positive') &
550. (data_1['PR_STATUS'] == 'Positive') &
551. (data_1['HER2_STATUS'] == 'Negative')
552. )
553. clin_sub = data_1[hrp_her2n_mask].copy()
555. # 2. Further filter for HISTOLOGICAL_SUBTYPE == 'Lobular' or 'Ductal/NST'
556. supported_subtypes = ['Lobular', 'Ductal/NST']
557. clin_sub = clin_sub[clin_sub['HISTOLOGICAL_SUBTYPE'].isin(supported_subtypes)]
559. # 3. Extract/merge PDCD1 and CD274 expression
560. expr_df = data_0[data_0['Hugo_Symbol'].isin(['PDCD1', 'CD274'])]
561. expr_long = expr_df.set_index('Hugo_Symbol').T
562. expr_long.index.name = 'PATIENT_ID'
563. expr_long = expr_long.reset_index()
564. expr_long['PDCD1'] = pd.to_numeric(expr_long['PDCD1'], errors='coerce')
565. expr_long['CD274'] = pd.to_numeric(expr_long['CD274'], errors='coerce')
567. # 4. Merge
568. merged = pd.merge(
569. clin_sub[['PATIENT_ID', 'HISTOLOGICAL_SUBTYPE']],
570. expr_long[['PATIENT_ID', 'PDCD1', 'CD274']],
571. on='PATIENT_ID', how='inner')
573. # Prepare results dict for summaries
574. results = {}
575. plots = []
576. **for** gene **in** ['PDCD1', 'CD274']:
577. # drop missing
578. subset = merged.dropna(subset=[gene, 'HISTOLOGICAL_SUBTYPE'])
579. group_ILC = subset[subset['HISTOLOGICAL_SUBTYPE'] == 'Lobular'][gene]
580. group_IDC = subset[subset['HISTOLOGICAL_SUBTYPE'] == 'Ductal/NST'][gene]
581. # Check normality for each group
582. p_normal_ILC = shapiro(group_ILC)[1] **if** len(group_ILC) >= 3 **else** np.nan
583. p_normal_IDC = shapiro(group_IDC)[1] **if** len(group_IDC) >= 3 **else** np.nan
584. # Mann-Whitney or t-test
585. **if** (p_normal_ILC > 0.05) **and** (p_normal_IDC > 0.05):
586. stat, pval = ttest_ind(group_ILC, group_IDC, nan_policy='omit')
587. test_type = 't-test'
588. **else**:
589. stat, pval = mannwhitneyu(group_ILC, group_IDC, alternative='two-sided')
590. test_type = 'Mann-Whitney U'
591. results[gene] = {
592. 'n_ILC': len(group_ILC), 'median_ILC': np.median(group_ILC),
593. 'n_IDC': len(group_IDC), 'median_IDC': np.median(group_IDC),
594. 'test_type': test_type, 'statistic': stat, 'p_value': pval
595. }
596. # boxplot
597. plt.figure(figsize=(7,5))
598. sns.boxplot(x='HISTOLOGICAL_SUBTYPE', y=gene, data=subset, order=['Ductal/NST', 'Lobular'])
599. plt.title(f'{gene} expression: HR+HER2- only')
600. plt.ylabel(f'{gene} expression')
601. plt.xlabel('Histological subtype')
602. plots.append(plt.show())
604. print('RESULTS (ILC vs IDC, HR+HER2- only):')
605. **for** gene, res **in** results.items():
606. print(f"{gene}: n_ILC={res['n_ILC']} median_ILC={res['median_ILC']:.3f} | n_IDC={res['n_IDC']} median_IDC={res['median_IDC']:.3f} |

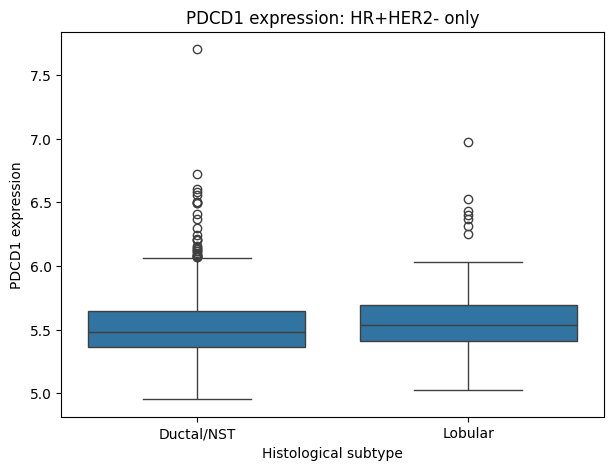

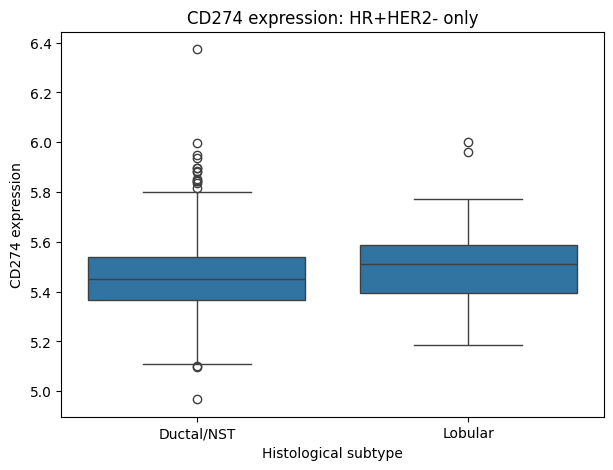

**multiplex immunofluorescence**

A total of 21 formalin‑fixed, paraffin‑embedded (FFPE) breast tissue sections were analyzed, including 9 non‑diseased samples and 12 ILC samples (2 triple‑negative, 4 HER2‑positive, and 6 hormone receptor–positive/HER2‑negative). Slides were baked at 60°C for 1 hour, deparaffinized in xylene, and heat‑induced antigen retrieval using an Epredia PT Module (Epredia, Kalamazoo, MI). Following antigen retrieval, slides were washed three times in Lunaphore multistaining buffer (5 min per wash) and subsequently loaded onto the Lunaphore COMET automated sequential immunofluorescence platform (Lunaphore Technologies, Tolochenaz, Switzerland).

For each sample, a 12.5 mm × 12.5 mm imaging area was aligned under the COMET microfluidic slide cover on the instrument’s imaging stage. An initial autofluorescence image of the imaging area was acquired, followed by 15 iterative cycles of immunofluorescent staining, image acquisition, and signal elution, all performed automatically by the COMET system. In each cycle, up to two primary antibodies (one rabbit-derived and one mouse-derived) were applied simultaneously and then detected with the appropriate Alexa Fluor–conjugated secondary antibodies specific to each primary’s host species. The primary antibodies include CD4 clone D6N8B rabbit 1:100 (Cell Signal technology), Cytokeratin clone AE1/AE3 mouse 1:150 (Agilent), CD3 MRQ-39 rabbit 1:1000 (Sigma-Aldrich), CD8 clone 4B11 mouse 1:100 (Bio-Rad), PD1 EPR4887 rabbit 1:400 (Abcam) and PD-L1 clone IHC411 rabbit 1:150 (Genome Me). The secondary antibodies include DAPI 1:1000, goat anti-mouse IgG Alexa fluor 647, goat anti-rabbit IgG Alexa fluor 647, and goat anti-mouse IgG Alexa Fluor 655 (Thermo Scientific, Swedesboro NJ). Fluorescence image acquisition was carried out at 20× magnification. After completion of all cycles, the instrument automatically compiled a stacked multi‐channel OME‐TIFF file encompassing all fluorescence channels.

The multi-channel OME-TIFF image was then exported via Horizon Viewer software after applying background subtraction. The images were imported into the HALO AI digital pathology software (version 4.2, Indica Labs, Albuquerque, NM) for visualization and quantitative image analysis. Using HALO AI’s trainable deep-learning module (MiniNet), two convolutional neural network classifiers were developed: one to identify and mask imaging artifacts, and another to segment tissue into epithelial versus stromal regions. Cell nuclei were segmented in the DAPI channel using HALO’s Nuclei Segmentation v2 algorithm, with a 3 μm cytoplasmic expansion radius applied to define whole-cell boundaries. Cell phenotypes were assigned by applying intensity thresholding to nuclear and cytoplasmic signals for each biomarker, with a minimum percentage of marker-positive area (“percent completeness”) required for a cell to be considered positive. The cell counts were The cell counts were region-of-interest based and normalized to tissue area. All statistical analyses were performed using GraphPad Prism 10 (GraphPad Software, San Diego, CA). Analysis Kruskal-Wallis test with Dunns correction for multiple comparisons.
